# Synthetic epigenomes for open cross-platform episignature classifiers

**DOI:** 10.1101/2025.03.28.25324859

**Authors:** Federico Ferraro, Livija Bardina, Mark Drost, Herma C. van der Linde, Michelle de Groot, Fatemeh Peymani, Amandine Santini, Helena Rasche, Mathis Hildonen, Erika Souche, Sanaa Choufani, Natalie Chaddock, Angele May, Daphne Smits, Prajkta Kallurkar, Bianca M. de Graaf, Rachel Schot, Erwin Brosens, Yolande van Bever, Alice Brooks, Laura Donker Kaat, Ilse Groeneweg - van der Heiden, Merel O. Mol, Nienke van Engelen, Anna L. de Boer, Natalie J. Wood, Epi2Diag collaborators, Ruizhi Deng, Tahsin Stefan Barakat, Wouter P. te Rijdt, Judith M.A. Verhagen, Virginie J.M. Verhoeven, Natalja Bannink, Ieva Grinfelde, Marija Jurcenko, Ieva Micule, Madara Redele, Marija Rozevska, Gita Taurina, Lenka Noskova, Christopher Campbell, David Gokhale, Ingo Kurth, Axel Weber, Michael Zech, Matias Wagner, Siddharth Banka, Rosanna Weksberg, Zeynep Tümer, Joris Vermeesch, Camille Charbonnier, Gijs Santen, Holger Prokisch, Florian Kraft, Christian Gilissen, Tjakko J. van Ham, Tjitske Kleefstra, Dmitrijs Rots

## Abstract

DNA methylation (DNAm) signature (episignature) classifiers support diagnosis of rare diseases. Yet episignature testing is confined to a dedicated array platform, and most classifiers are neither openly available nor fully reproducible. Laboratories cannot train their own classifiers as positive controls are scarce and cannot be shared under privacy regulations. Here, we develop and release MethaDory, a framework to train episignature classifiers using synthetic DNAm profiles generated in silico from control data and summary statistics of affected individuals from any platform of interest. We trained 188 classifiers spanning 102 rare disorders and validated 154 across 915 individuals with an episignature-associated diagnosis, 533 healthy controls profiled with DNAm arrays, Oxford Nanopore, and PacBio sequencing, together with 7 international centres. MethaDory achieved an average F1 0.9 on arrays, matching or outperforming existing approaches. We show MethaDory classifier application to long-read genome data, facilitating diagnoses in unsolved cases and in ultra-rapid diagnostic settings.

## Introduction

About 30-50% of individuals with suspected rare genetic diseases remain undiagnosed after first-line testing by short-read exome and genome sequencing^1–6^. A promising layer that could increase diagnostic yield is the epigenome. In 2013, it was observed that some monogenic developmental disorders (DDs) are associated with genome-wide, disorder-specific DNA methylation (DNAm) alterations in blood^7^. These alterations, referred to as DNAm signatures or episignatures, have been used to build machine learning classifiers and have improved the diagnostics of DDs by supporting the interpretation of variants of uncertain significance (VUS)^8,9^ and by guiding molecular diagnostics in DNA-testing “negative” individuals^8–12^. Moreover, episignatures have also been used to study the underlying molecular mechanisms of diseases and to define new disease entities^13–18^. While episignature testing increases the diagnostic yield, it is generally performed on dedicated DNAm arrays^19^ and is usually requested after sequencing was performed and identified a VUS in genes with an episignature or in unsolved cases with relevant phenotype. This sequential testing increases the diagnostic odyssey and costs. Long-read genome sequencing, e.g., by Oxford Nanopore Technologies (ONT) and Pacific Biosciences (PacBio), offers a much faster way to consolidate the detection of sequence variants and episignatures in a single assay^20^.

Reaching sample sizes sufficient for developing and validating episignature classifiers demands (inter)national collaborations, can take years, and is hindered by tightening privacy regulations in this field. Most published episignature classifiers are not publicly available or published with too little data for replication and validation. As a result, most diagnostic and research laboratories cannot adopt the episignature testing locally, cannot apply them to long-read sequencing data and must maintain a separate testing technology (DNAm arrays). To overcome the lack of individual-level data, a previous study trained DNAm classifiers using the averaged methylation profiles of affected and unaffected individuals^21^, which were available for a limited number of episignatures. This approach demonstrated that a reduced representation of the disorders’ DNAm profiles can be sufficiently informative to build episignature classifiers. However, this approach is at risk of overfitting to the training set due to lack of variability in the averaged profiles^22^.

Synthetic data strategies have previously been successfully applied to train classifiers in other domains where training data are scarce. These included medical imaging data (MRI, echocardiography, histology)^23^, text data (biomedical records)^24^, transcriptomic data^25^, and DNA methylation data for brain tumour classification^26^, but not for rare genetic diseases. In this study, we address the gaps in rare disease episignature testing by developing MethaDory, an approach to train cross-platform episignature classifiers using synthetic *in silico* epigenomes of affected individuals which mimic the epigenomes of patient samples, without requiring direct access to individual patient data. Because we build the synthetic epigenomes from control data generated on different platforms, the same published episignatures can even be translated from, e.g., arrays to long-read sequencing. We extemded this approach across 188 episignatures for 102 disorders using DNAm array and long-read data, benchmarked it against existing open alternatives, performed a multicentre international validation, together with an in-house clinical validation. We further showcase the power of utilizing long-read sequencing to simultaneously detect both genetic variants and episignatures to resolve previously unsolved cases, and in ultrarapid diagnostic settings. Finally, we release the DNAm classifiers with an open application to run these models locally.

In summary, we have built an open access route to episignature testing, available locally that does not require sharing patient data, and is durable across the long-read sequencing platforms, allowing the detection of episignatures together with genetic variants.

## Materials and methods

### Array DNAm data collection from publicly available sources and preprocessing

Eighty-five Gene Expression Omnibus (GEO) open datasets^8,27–105^, available as of July 15^th^ 2025 (**Supplementary Table 1**), were downloaded based on the pre-specified criteria: blood-derived raw DNAm array data (.idat files) for at least 6 healthy (control) individuals; or DNAm data of affected individuals with rare diseases with established episignatures. We only included studies using the Infinium HumanMethylation450 BeadChip, Infinium MethylationEPIC v1.0 BeadChip, or MethylationEPIC v2.0 BeadChip. Samples represent various ethnicities and age groups, and were stratified according to previously reported age groups^106^ (**Supplementary Figure 1**).

Array data analysis was performed in R v.4.5.1. Data were imported using minfi package v.1.50^107^, and underwent extensive quality control (QC) per study. WateRmelon v.2.10.0^108^ was used to predict sex-chromosome composition and age; blood and non-blood cell proportion deconvolution was performed with EpiDISH v.2.20.1^109^; and (genetically) identical samples, and potential contamination were detected with Ewastools v1.7.2^110^. Samples that failed the recommended Illumina quality metrics (Ewastools), with detected contaminations or mismatched for the reported sex, genetically identical samples, non-blood samples, and samples with a difference between predicted and reported age >10 years were removed. Additionally, samples from individuals >80 years of age were excluded because of the high prevalence of clonal hematopoiesis^111^. When multiple related samples were reported in a dataset, we included only the youngest individual. Next, cohort-level outliers were detected on principal component analysis (PCA), performed on the top 1% most variable autosomal CpGs using PCAtools v.2.16.0^112^. Array-type, age group, sex-stratified samples with values >3 standard deviations away from the average values of the first or second principal component were removed.

The signal intensities were normalized with normal-exponential convolution using out-of-band probes (Noob)^113^ with ChAMP v.2.34.0^114^. Study-specific single nucleotide polymorphisms were detected with MethylToSNP v.0.99.0^115^. Missing probe beta values were imputed using methyLImp2 v1.0.0^116^. For a few studies reporting affected individuals without the raw .idat files, we used the available processed data for testing purposes only but not training.

In total 5,328 unique control individuals out of 10,944 samples were retained (**Supplementary Figure 1**). The controls were randomly split into three subsets, while balancing for array-type, age groups, and sample sex: 5,000 samples were utilized for the training of the classifiers and generation of the synthetic epigenomes; 164 samples were used for the optimization of the models; 164 controls were assigned to the final testing of the models.

### Long-read sequencing data generation and preprocessing

The DNAm profiles of controls assessed with ONT or PacBio sequencing were obtained from individuals unlikely to have altered DNAm profiles from Erasmus Medical Center (Rotterdam, the Netherlands, Erasmus MC; 31 ONT controls), Radboud University Medical Center (Nijmegen, the Netherlands, Radboudumc; 402 PacBio controls), or Uniklinik RWTH Aachen (Aachen, Germany; 131 ONT controls). We included only individuals not affected by neurodevelopmental disorders, congenital anomalies, metabolic disorders, growth abnormalities, nor haematological disorders. Long-read DNAm profiles underwent quality control per center, as described above for the DNAm array data.

For ONT sequencing at Erasmus MC, genomic DNA from whole blood was fragmented, short fragments were removed, and processed according to ONT (Oxford, United Kingdom) ligation Sequencing Kit SQK-LSK114 protocol, and sequenced on a PromethION Flow Cells R10.4.1. ONT cohort had an average read depth of 38x. Basecalling with modification, and alignment to the reference genome (GRCh38) were performed with dorado v. 1.0.0, using the basecalling models hac@v5.0.0,5mCG_5hmCG or sup@v5.0.0,5mCG_5hmCG or dorado v 2.0.0 and models hac@v6.0.0,5mCG_5hmCG. DNA methylation was summarized with modkit v0.5.0 or modkit v0.6.2 and reformatted. PacBio sequencing from Radboudumc data was generated between January 2024 and January 2025 on PacBio Revio (Pacific Biosciences) using a single SMRT cell per sample to obtain 24-hour movies, to obtain 30X average genome coverage at 15-18kb read length. Preprocessing was previously described in Ivashchenko et al.^117^ read alignment was performed with PBMM2, methylation calls were performed with primrose version 1.0.0, and then tabulated with pb-cpg-tools version 2.3.1. CpG sites at SNPs (>1% MAF in gnomAD v4 ancestral populations) were removed.

PacBio data from Erasmus MC, genomic DNA from blood was fragmented, short fragments were removed and processed according to PacBio HiFi prep kit 96 protocol. The library was sequenced on a PacBio Revio using a single SMRT Cell and sequenced for 24 hours to obtain read depth of 30x. Methylation calling was performed on Revio instrument with Jasmine v.2.3.0. Sequencing data was aligned with minimap2 v.2.30 and DNA methylation was summarized with pb-CpG-tools run in ‘default’ mode.

### ONT downsampling experiments

Downsampled DNAm data replicates of the ONT test set were generated similarly to downsample_methylKit.py (github: nebiolabs/methylation_tools). Reads were independently sampled with probability p = target coverage / mean(valid coverage), with valid coverage being (Nmod + Ncanonical +Nother_mod), based on modkit specifications. All seven per-site count fields were thinned jointly and valid coverage and percent modified recomputed. Three independent draws of each target read depth (5x, 10x, 15x, 20x and 30x) were performed.

### Generation strategy of synthetic epigenomes of affected individuals and mosaics

We generated synthetic epigenomes of affected individuals for each disorder for which an episignature or profile could be extracted from literature. We use the term “*synthetic* epigenomes of affected individuals” to distinguish this synthetic data from actual DNAm data of “affected individuals”, i.e. patients with a confirmed clinical and molecular diagnosis (collected from literature, in house dataset, and international collaborations).

For each synthetic epigenome (SE) of a target condition and episignature, we approximated the methylation level (β) data distribution at a specific CpG site *i* as:

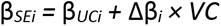

with β*UCi* being the starting methylation value of the unaffected individual (UI) from the control dataset used to generate the SE, *Δ*β*i* the average difference between affected and unaffected individuals reported by the studies as part of the target episignature, and VC - the modelled variability coefficient, which captures the variability of the effect sizes. Since the real distribution of the VC in episignatures is not known, we adopted a heuristic solution and tested sampling the VC from beta distribution *Beta*(α, β), where α and β are positive shape parameters. For all the distributions, we centered the mode of the VC at the published effect size. Distribution parameters and detailed performance is reported in **Supplementary Table 2.**

We also generated synthetic mosaic cases, but have used affected individuals with clear haploinsufficiency-causing variants following Oexle et al.^118^. Briefly, mosaics were simulated over a span of variant allele frequencies (VAF) of 0.1-0.45, with steps of 0.05, as weighted average (by the allele frequency of interest) of a typically presenting affected individual and one of 25 random unaffected individuals from the set of controls used for the optimization of the models.

### Episignature CpG site quality control and harmonization

As different studies adopt variable strategies to deal with CpG-level QC^119^, we have harmonized CpGs episignature site selection. Since the published signatures have been derived using different DNAm array platforms (Infinium HumanMethylation450 BeadChip or Infinium MethylationEPIC v1.0 or v2.0), we retained probes present in the MethylationEPIC v1.0 and v2.0 arrays. Then, we filtered out non-specific probes, probes prone to batch effects, and probes on the sex chromosomes. Probes were removed also if overlapping variants were detected by MethylToSNP v.0.99.0 with global allele frequency >5% and confidence score >0.5 in at least one of the control datasets; probes mapping to copy number variant (CNV) loci included in the corresponding CNV signatures were also removed. We also removed sites that were found among the top 1% most variable in the control database and sites having an effect size at the limit of the episignature resolution by the arrays |Δβ|≤0.05. We also removed sites with Pearson’s correlation ≥ 0.9 with age in controls. Finally, for signatures with >500 reported sites passing the QC, probe redundancy was reduced by removing CpG-sites with correlation ≥ 90% within each chromosome and then selected the top 500 with the highest calculated -|Δβ| * log10(p-value), expecting these to be the most informative and having the highest confidence^13^.

### DNAm classifiers development and testing

We trained and benchmarked 10 different machine-learning (ML) models using caret v.6.0- 94a^120^: support vector machines (SVM) with a linear kernel ( ‘svmLinear1’ and ‘svmLinear2’), with radial basis function kernel (method ‘svmRadialSigma’), and 8 other models selected to encompass linear and non-linear methods: nearest shrunken centroids (‘pam’), linear discriminant analysis (‘lda’), quadratic discriminant analysis (‘qda’), elastic net regularized generalized linear models (‘glmnet’), random forest (‘ranger’), stochastic gradient boosting (‘gbm’), neural network (‘nnet’), and the naïve Bayes (‘naive_bayes’). Hyperparameter tuning was performed per episignature using grid search within 5-fold cross-validation.

All the models were set to ‘probability’ to generate scores in the interval 0-1, with higher values representing a “positive” classification for the episignature.

The final neural-network (NN) classifiers were trained in PyTorch (v.2.5), to enable GPU acceleration and Bayesian hyperparameter optimisation in Optuna v.4.8^121^ to select learning rate and number of neurons. For each episignature, the networks were trained on the same *in silico* data used for the SVMs. Each NN consisted of a single fully connected hidden layer with sigmoid activation and a single-unit output. NNs were optimised with Adam^122^ by minimising the binary cross-entropy loss for up to 500 epochs, with early stopping after 50 epochs without improvement of the validation loss.

### MethaDory classifiers final architecture

The final MethaDory classifiers were built by combining synthetic epigenomes, generated *in silico* from control data, with real controls to form the training set, not overlapping with the samples used for the validation or testing of the models. On this set, we trained two complementary models for each episignature: a linear SVM and a feed-forward NN, selected based on our benchmarking (**Supplementary Figure 6**). To make the predictions more robust, we split the controls into three stratified folds and used each of the three fold pairs twice, swapping which fold was used to generate the synthetic cases and which served as controls. This produced six training sets, and, therefore, six SVM and six NN models, per episignature. Each sample was then scored by all twelve models. Within each model type we discarded the highest and lowest probabilities and averaged the rest, and the final MethaDory score is the mean of the SVM and NN trimmed means. Scores were reported per episignature as a probability between 0 and 1. The same procedure was applied separately to DNAm arrays, ONT, and PacBio data.

### Head-to-head comparison of MethaDory with EpigenCentral and NSBEpi

Currently, two public DNAm classification approaches are available. First, EpigenCentral^123^, hosted by The Hospital for Sick Children, Toronto, is a freely accessible web portal for classification analysis of episignatures. The models included in this platform were trained using affected individuals and matched unaffected individual data. For each technology, we uploaded the pre-processed beta matrices, before missing value imputation, and EpigenCentral was run selecting the corresponding technology model option. For EpigenCentral and MethaDory we used a threshold of 0.5 to call a sample “positive”. We used the classifier threshold of 0.5 as a criterion for computing binary performance metrics, but the authors of EpigenCentral treat scores between 0.4 and 0.6 as intermediate rather than assigning them to either cases or controls.

Second, NSBEpi, developed by Geysens et al.^21^, 2025, using only methylation average beta values of individuals affected from 34 disorders and unaffected individuals^124^. NSBEpi consists of a linear SVM for each disorder, and it reports for each tested sample the most similar episignature among the 34. NSBEpi does not report a probability score, and we used the threshold of 0.35, as recommended by the authors.

### Evaluation metrics

To assess the performance of the DNAm classifiers, we employed: precision, the proportion of correctly predicted positives out of all predicted positive results; sensitivity (also called recall or true positive rate), the proportion of correctly predicted positive results out of all actual positive results; F1 score, the harmonic mean of precision and sensitivity.

### Published episignature stability analysis using synthetic epigenomes

To evaluate the discriminative capacity of each episignature, we used a bootstrapped simulation framework with 1,000 iterations. In each iteration, 60 control samples were randomly selected without replacement from the available pool and split into two equal groups of 30. One group was retained as controls, while the other was used to generate synthetic cases as described above. For each iteration and each episignature, principal component analysis was performed on the combined matrix of synthetic cases and controls using all CpG sites included in the episignature. The first two principal components were used to quantify separation between case and control groups using four clustering metrics: mean silhouette width, Calinski-Harabasz index, Davies-Bouldin index, and Dunn index. Silhouette widths were computed using Euclidean distance with the R package cluster v.2.1.8.1, and the remaining indices were calculated using the clusterCrit package v.1.0.3^125^.

### Concordance of observed and reported episignature effects

For each signature with at least two non-mosaic affected individuals in the DNAm array test set, the reported Δβ was compared with the observed effect, i.e. the mean deviation of the affected individuals from the mean β of the test controls at each episignature CpG site. Effect recovery was defined as the slope of the least-squares regression of the observed effect on the reported Δβ, with a value of 1 corresponding to the full reported effect and 0 to no effect. Pearson’s correlation between observed and reported effects was used to assess whether the reported pattern was reproduced irrespective of its magnitude. To estimate the correlation expected by chance, n test controls (n, number of affected individuals for that signature) were randomly drawn as pseudo- cases in each of 1,000 iterations and compared with the remaining controls. One-sided empirical p-values ((k + 1)/(B + 1), with B = 1,000 iterations, and k the iterations reaching at least the observed correlation) were corrected with the Benjamini-Hochberg method. The association of effect recovery with classifier F1 and with the difference between the mean scores of affected individuals and controls was tested with Spearman’s correlation.

## Results

### Synthetic epigenomes can replace patient data to train robust episignature classifiers

We harmonized DNAm data for almost 6,000 individuals spanning controls and individuals affected by rare disease for the following analyses (**Figure 1**).

**Figure 1.**
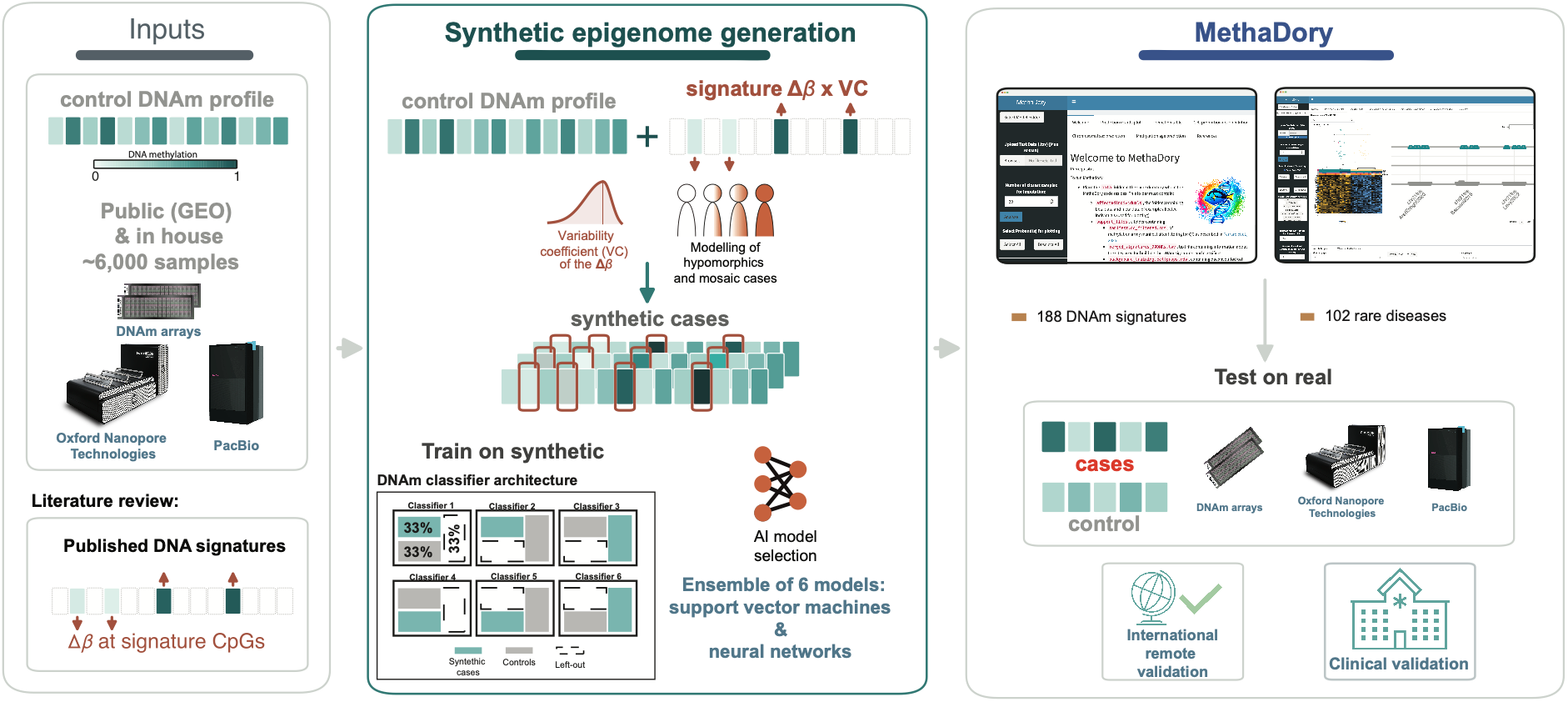
Overview of the MethaDory framework: synthetic epigenomes enable cross-platform DNAm classifiers for developmental disorders and congenital anomalies without patient samples. Control DNA methylation (DNAm) profiles were assembled from publicly available and in-house sources across three platforms (methylation arrays, Oxford Nanopore Technology (ONT), and PacBio long-read genomes) together with published DNAm signatures (signature CpG sites and their reported case-versus-control effect sizes) and a curated set of molecularly diagnosed affected individuals used only for evaluation. For each disorder, in-silico synthetic cases are generated by adding the published signature effect size, scaled by a variability coefficient, to the methylation values of unaffected control samples. Train-on-synthetic-test-on-real (TSTR) strategy and final classifier design. Classifiers are trained on synthetic cases and unaffected controls and evaluated on real affected individuals and controls held out from training. To stabilize predictions, the final design splits the controls into three folds and trains six models on pairwise fold combinations, reporting the trimmed mean and standard deviation of the six scores. All resulting classifiers are released in MethaDory, a free, open-access application that runs locally and requires no transfer of patient data and was validated internationally remotely and in-house.

First, to evaluate whether synthetic epigenomes of affected individuals can be recognised as cases by episignature classifiers trained on real individuals, we generated 1,800 synthetic epigenomes using 100 random controls for 18 episignatures (available by February 2025) from EpigenCentral, which provides classifiers trained on real affected individuals and controls^123^. The EpigenCentral classifiers distinguished the synthetic epigenomes as affected cases from the original 100 controls with high accuracy (average F1 = 0.95, **Supplementary Figure 2, Supplementary Table 3**). We, therefore, hypothesized that synthetic epigenomes could also be used for the training of the episignature classifiers. Next, we trained SVM DNAm classifiers for 14 disorders using solely synthetic epigenomes and controls, focusing on disorders for which we had ≥10 affected individuals available, and a classifier also present in EpigenCentral (**Supplementary Figure 3**). To select the optimal number of training samples, we trained classifiers with an increasing number of synthetic epigenomes and controls (from 25 to 1500 of each stratum, **Supplementary Figure 4**) and evaluated them on 213 real affected and 164 unaffected individuals. We observed that increasing synthetic epigenome training-set size improved case-control separation, and generally reduced prediction variability across the trained models. This behavior is also seen, when training on real samples (**Supplementary Figures 4-5, Supplementary Tables 4-5**), and, therefore, we decided to use the maximum number of available samples. This training approach extended to other machine learning (ML) models, with NNs achieving the highest performance of the 10 tested models (F1 0.992 **Supplementary Figure 6, Supplementary Table 6**). As a heuristic choice the top two F1 models were used in following experiments: the linear SVM, widely employed^119^, and the NN that rescued the performance of the chr16p11.2 deletion^13^ signature.

To increase classifier sensitivity to intermediate effect (hypomorphic) and mosaic variants, we modelled the synthetic epigenomes of individuals mosaic for pathogenic variants by including a coefficient of variability (VC) in the synthetic generation formula. We observed improved sensitivity for synthetic mosaics for both SVMs and NNs (delta average sensitivity +0.10, exact binomial sign test p<10-12), while SVMs with VCs also show higher detection sensitivity than EpigenCentral models trained directly on affected individuals (delta average sensitivity +0.14, exact binomial sign test p=0.002, **Supplementary Figure 7, Supplementary Table 7**). Considering the above, in the final architecture, we score each sample with an ensemble of six SVMs and six NN models trained on complementary folds, using a robust ensemble average of these two model types as the final prediction (**Methods**).

Finally, after demonstrating that the synthetic epigenome can provide robust classifier training input, we expanded the episignature training for 188 blood-derived curated episignatures spanning 102 rare disorders and clinical syndromes (93 genes and genomic loci, two prenatal exposures, and one recurrent constellation of embryonic malformations, RCEM) reported in 81 studies^7,8,13–18,85–87,95,96,98,102–105,124,126–186^ (**Supplementary Table 8**). We trained MethaDory-array classifiers using 5,000 controls and MethaDory-long-read classifiers using ONT data from 162 controls and PacBio data from 396 controls.

### MethaDory classifiers show high accuracy on DNAm arrays

We first evaluated the MethaDory DNAm classifiers trained on synthetic array-based epigenomes on a test set of array-based data from 241 affected individuals (143 signatures in 73 disorders) and 164 controls excluded from the model training. Overall, the MethaDory DNAm classifiers obtained an average F1 of 0.90, including signatures with “moderate effect sizes”, i.e., those published with limited predictive power (e.g., *ANKRD11*^148^*, KMT2C*^14^) and discordant signatures for the same disorder (e.g., *CHD8*^124^) (**Figure 2, Supplementary Table 9, 10**). MethaDory models scored the four mosaic individuals in our cohort (NM_006015.6(*ARID1A*):c.666C>A p.(Tyr222Ter) VAF ∼19%, chr16p11.2 deletion, affected, VAF ∼12.5% blood, NM_022552.5*(DNMT3A*):c.2312G>A p.(Arg771Gln), unaffected, VAF ∼50% blood, VAF 0% saliva, VAF ∼10% urine^98,187^, and NM_024757.5(EHMT1):c.2273T>C p.(Leu758Pro), VAF ∼6% blood) higher than unaffected individuals.

**Figure 2.**
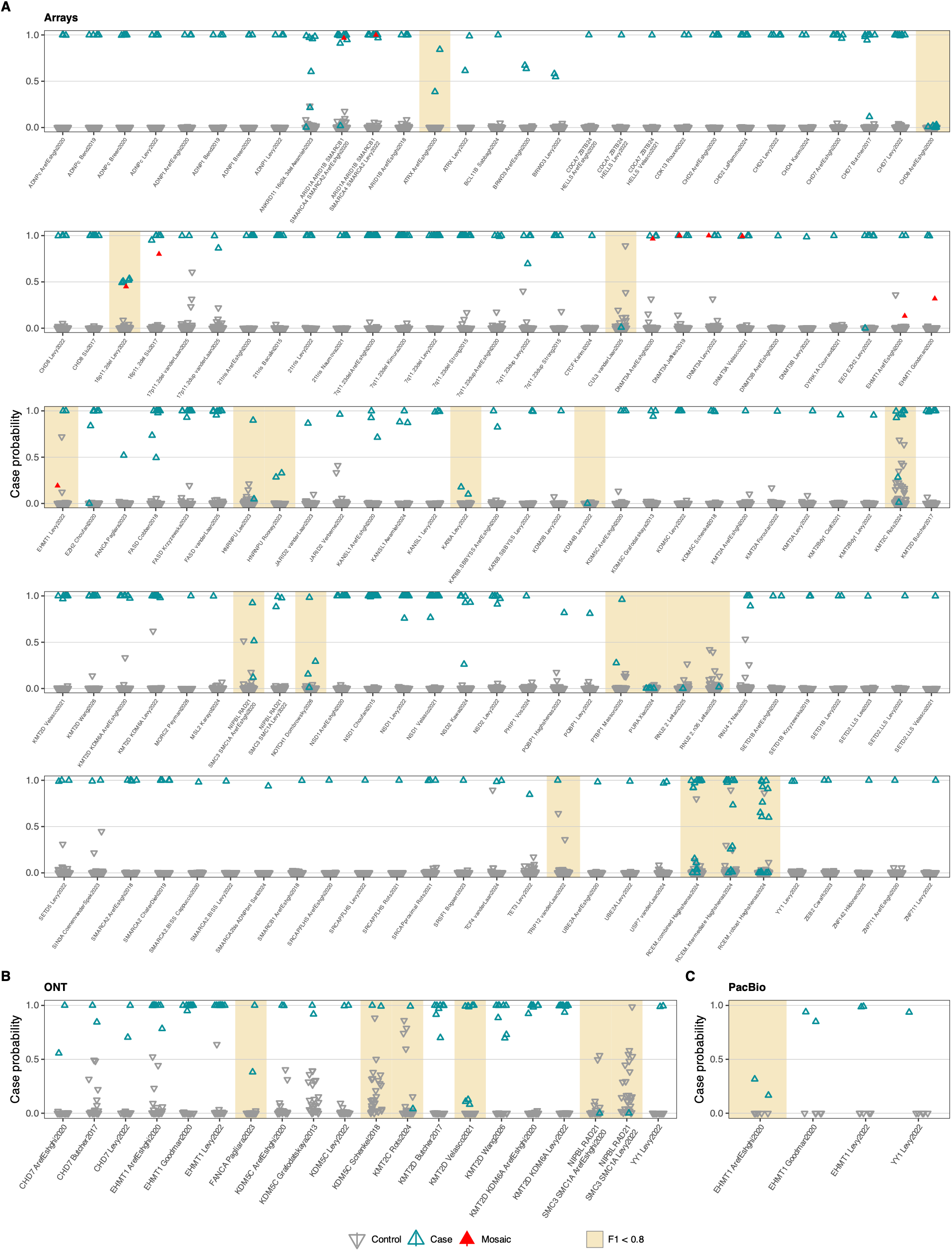
Array-trained MethaDory classifiers identify affected individuals from controls across technologies. **A)** MethaDory array-trained classifiers evaluated on 241 samples from affected individuals (spanning 143 signatures) and 1C4 controls. **B)** MethaDory array-trained classifiers applied directly to DNAm profiles derived from 57 ONT long-read genomes (2S affected individuals and 28 controls). **C)** MethaDory array-trained classifiers applied directly to 8 PacBio long-read genomes (3 affected individuals and 5 controls). Each point represents the prediction score of a sample for the indicated DNAm signature; whiskers represent the scores obtained by the neural network and SVM. Higher scores indicate a classification as of case.

Since the synthetic epigenomes are generated from the published effect sizes, a classifier can only perform well if the published episignature replicates in new individuals. To distinguish whether the lower performance of some of our classifiers was due to our training approach or to the published episignatures themselves, we tested whether the affected individuals in our test set reproduced the effects independently of any classifier. For 99 signatures with ≥2 non-mosaic test cases (210 individuals), we compared observed case-control differences with published effect sizes. Cases recovered a median of 91% of the published effect, but 11 signatures recovered less than half (**Methods, Supplementary Figure 8, Supplementary Table 8**). Effect recovery correlated with classifier performance (F1 Spearman’s ρ = 0.49): signatures recovering less than half of the effect achieved an average F1 of 0.50, compared with 0.98 for those recovering at least 80%. For the same seven CHD8 individuals, the Aref-Eshghi et al. signature^124^ recovered 15% and had a sensitivity of 0, whereas the Siu et al. signature^130^ recovered 93% and had a sensitivity of 1. F1 also correlated with four complementary case-control separation metrics computed on the training synthetic cases (**Methods, Supplementary Figure 9, Supplementary Table 8**). Together, these results suggest that lower-performing classifiers mostly reflect signatures whose published effects do not replicate in the independent affected individuals we had access to.

MethaDory classifiers were trained to distinguish one disorder from controls. Generation of one-versus-all classifiers is not possible with the current published data as they provide only selected probes per disorder, so the effect sizes of the other signatures at these loci are not known. We assessed the cross-disorder specificity of the signatures by counting the number of positive classifications (score ≥ 0.5, restricted to signatures with ≥ 2 cases) obtained by affected individual samples in signatures different from the one for the expected condition. Most diagnoses cross-reacted with a single signature, in several cases a paralog or same-complex gene, except for *NSD1,* which showed broad cross-reactivity (19 signatures) consistent with the large and broad methylation alteration in this disease^13,89,102^ (**Supplementary Table 11**).

### MethaDory DNAm classifiers can classify on long-read DNAm data

To evaluate performance on long-read data, we tested MethaDory classifiers on 57 ONT (29 affected individuals and 28 controls, over 9 disorders) and 8 PacBio (3 affected individuals and 5 controls, over 2 disorders) genomes. Both ONT and PacBio sequencing recovered all pathogenic DNA variants, correctly identifying 24 SNVs/indels and 5 structural variants (**Table 1**). Synthetic array epigenome-trained MethaDory classifiers correctly distinguished the affected individuals from controls, achieving an average F1 of 0.81 on ONT data and average F1 of 0.75 on PacBio data (**Figure 2B-C, Supplementary Table 8, 10**), showing that classifiers built from synthetic array epigenomes can also be applied to long-read platforms.

**Table 1:**
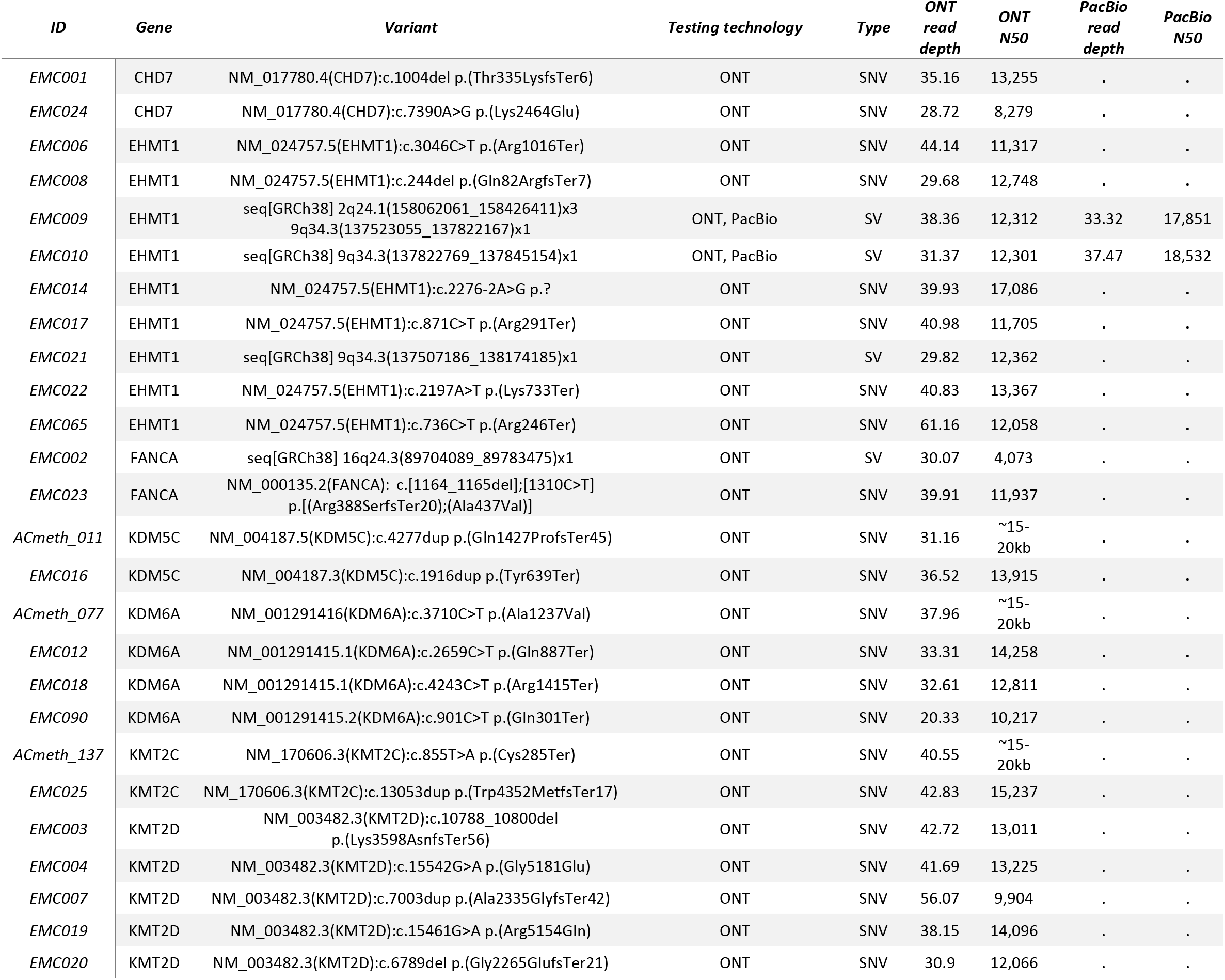
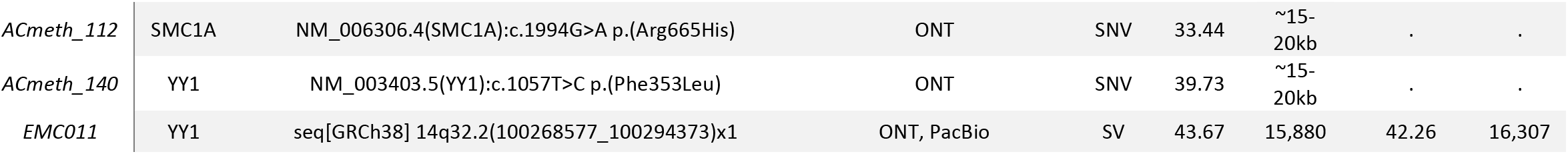
Overview of samples sequenced with long-read sequencing technologies and used for episignature testing.

Next, we tested the sequencing depth in ONT needed on the classifications by downsampling the ONT test set (**Methods**). We show that array-trained classifiers reached a plateau at 15x-20x (mean sensitivity 0.94, specificity 0.99), but ONT-trained classifiers improved more slowly (**Supplementary Figure 10**). These results suggest that array-trained classifiers can transfer to long-read data, requiring at least 20x of ONT sequencing data.

A subset of 13 individuals was assayed on arrays and at least one long-read platform (11 on DNAm arrays and ONT, 2 on arrays, ONT and PacBio; **Supplementary Figure 11, Supplementary Table 12**). For each sample and signature, the call was identical across the technologies used to assess the sample in 114/117, 112/117 and 115/117 comparisons with the array, ONT- and PacBio- trained classifiers, respectively. The *EHMT1* signature of Aref-Eshghi et al.^124^ scored one PacBio- measurement of a *EHMT1* individual below threshold under all three training sets, while the arrays and ONT measurements and the other two *EHMT1* signatures were positive.

We also assessed the impact of the training set size on ONT and PacBio (**Supplementary Figure 11**) and observed a similar improvement in performance with larger training set size also on long- read data (**Supplementary Figure 12**).

Finally, we tested the MethaDory long-read-trained models on long-read DNAm data. MethaDory-ONT classifiers achieved an average F1 of 0.78 on ONT test samples, and MethaDory- PacBio classifiers achieved an F1 of 0.75 on PacBio test samples (**Supplementary Figure 13, Supplementary Table 9-10**). The lower performance of the long-read models could be explained by the influence of the smaller number of control samples to generate synthetic epigenomes currently available.

### Comparison of the DNAm MethaDory classifiers with previous approaches

We next compared the performance of MethaDory array-based classifiers (given their largest training dataset size) with the ones in EpigenCentral and NSBEpi. We matched samples to the 10 disorders (CHARGE, *CHD8*, chr21 trisomy, Dup7, Williams, *EHMT1*, *KMT2D*, *NSD1*, *SRCAP*, *SMARCA2* syndromes) and signatures shared across all three classifiers and used 403 arrays (239 affected individuals and 164 controls), 47 ONT (19 affected individuals and 28 controls), and 7 PacBio (2 affected individuals and 5 controls) samples. MethaDory achieved comparable or superior average precision and sensitivity on arrays, ONT and PacBio data (**Table 2**, **Figure 3, Supplementary Tables 13-15**). Two DNAm array samples were classified as controls by all approaches despite expected to be cases. The first EMC031, carrying the mosaic NM_024757.5(*EHMT1*):c.2273T>C p.(Leu758Pro) variant had intermediate scores in MethaDory and EpigenCentral that might reflect the low mosaicism (∼6% VAF in blood), and the other was GSM3227780 carrying the NM_003482.3(*KMT2D*):c.11800C>T p.(Gln3934Ter)^97^.

**Figure 3.**
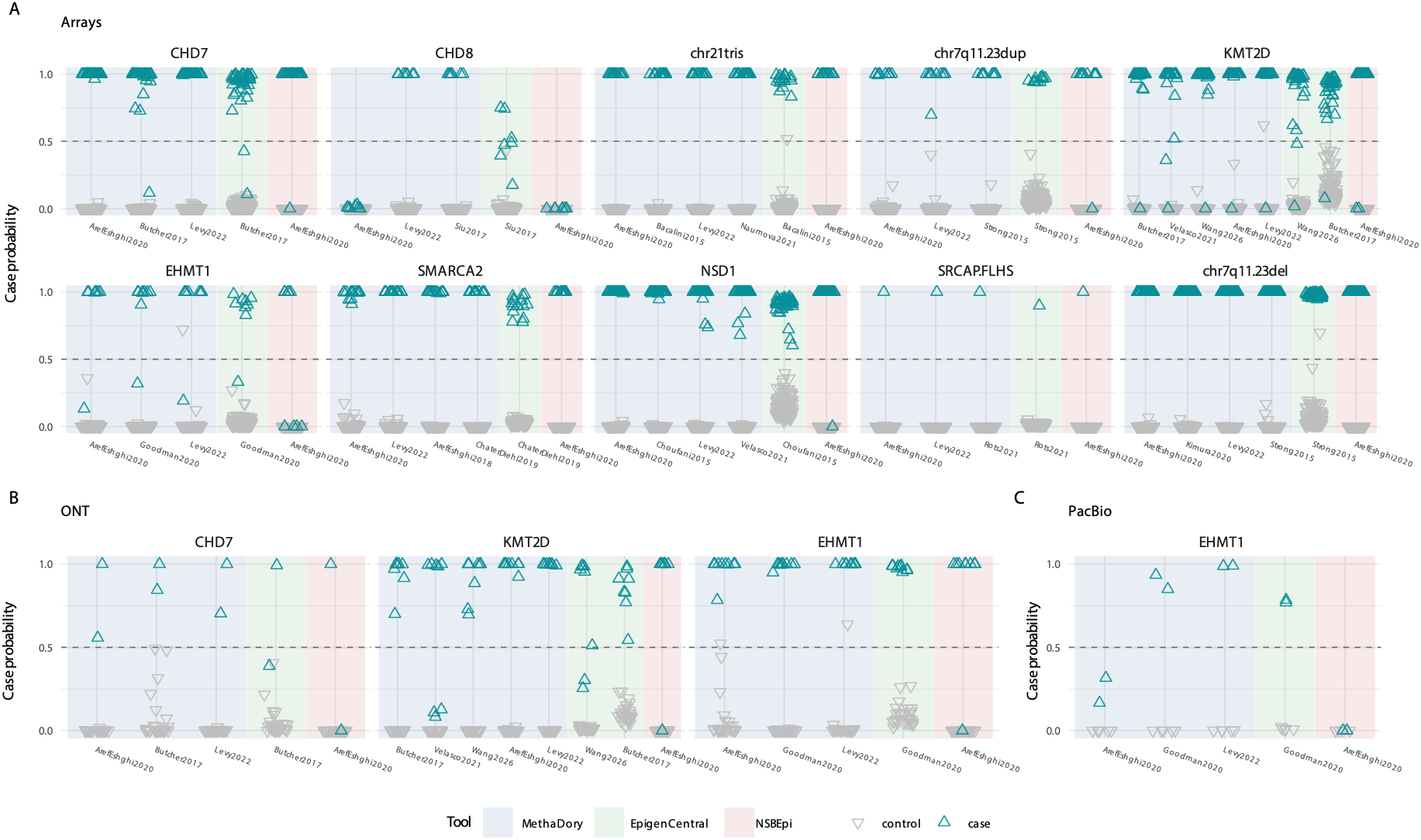
Comparison of MethaDory with EpigenCentral and NSBepi on the subset of samples that could be scored by all three approaches. **A)** 403 arrays (23S affected individuals and 1C4 controls), **B)** 47 ONT (1S affected individuals and 28 controls), and **C)** 7 PacBio (2 affected individuals and 5 controls). Since NSBepi does not output a probability score, predictions of “control” were assigned a value of 0 and predictions of “disease” a value of 1. Summary of classification metrics for the three approaches are reported in Table 2.

**Table 2:**
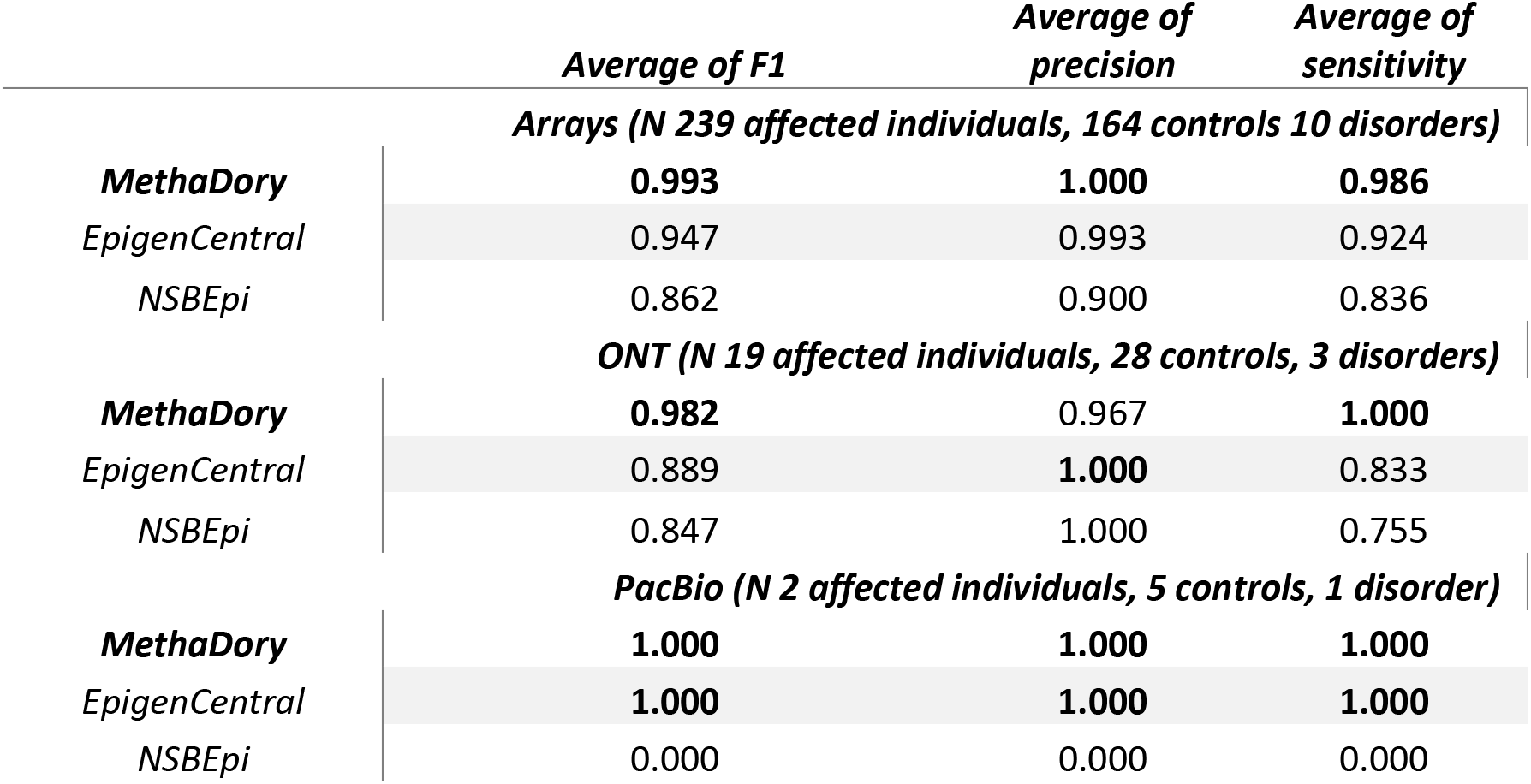
Classification performance of MethaDory, EpigenCentral, and NSBEpi on the same subset of samples. For MethaDory and EpigenCentral, we summarized predictions where multiple signatures are present with a majority vote.

|  | ***Average of F1*** | ***Average of precision*** | ***Average of sensitivity*** |
|---|---|---|---|
|  | ***Arrays (N 239 affected individuals, 164 controls 10 disorders)*** |  |  |
| ***MethaDory*** | **0.993** | **1.000** | **0.986** |
| *EpigenCentral* | 0.947 | 0.993 | 0.924 |
| *NSBEpi* | 0.862 | 0.900 | 0.836 |
|  | ***ONT (N 19 affected individuals, 28 controls, 3 disorders)*** |  |  |
| ***MethaDory*** | **0.982** | 0.967 | **1.000** |
| *EpigenCentral* | 0.889 | **1.000** | 0.833 |
| *NSBEpi* | 0.847 | 1.000 | 0.755 |
|  | ***PacBio (N 2 affected individuals, 5 controls, 1 disorder)*** |  |  |
| ***MethaDory*** | **1.000** | **1.000** | **1.000** |
| *EpigenCentral* | **1.000** | **1.000** | **1.000** |
| *NSBEpi* | 0.000 | 0.000 | 0.000 |

### MethaDory independent external and clinical validation

To facilitate the use of the MethaDory classifiers, we developed an open-access MethaDory companion app that streamlines episignature classification and interpretation, distributed with all the 188 episignatures for DNAm arrays, ONT and PacBio data [Zenodo]. MethaDory is computationally lightweight, runs on an ordinary laptop, and returns results within minutes, without sharing sensitive data. It reports classification scores, and generates dimensionality- reduction plots, aiding interpretation with visual representation as previously recommended^19^.

MethaDory was independently validated via network of international partners (n=7) on their affected individuals, unaffected controls, and individuals with a molecular diagnosis of a disorder without an available episignature. Altogether 1,338 additional samples were evaluated across DNAm arrays, ONT, and PacBio, spanning 100 signatures and 54 genes and genomic loci (**Supplementary Tables 16-17, Supplementary Figure 14**).

On DNAm arrays (611 cases, 292 controls), MethaDory classifiers obtained an average F1 of 0.90 (average precision 0.97 and sensitivity 0.87, over 87 episignatures). We also scored 356 samples from individuals affected by an episignature-negative disorder, i.e. disease control, and obtained an average F1 of 0.89 (average precision 0.95).

For ONT, of the 22 cases, the array-trained MethaDory classifiers detected the expected episignature in 19/22 cases, while the single control was correctly classified by all signatures. The 3 negative cases had previously been reported to be negative in both NSBEpi and EpiSign^TM^ ^21^. The technology-matched MethaDory-ONT classifiers identified the same 19 cases with at least one expected signature.

On PacBio (13 cases and 43 controls, 8 signatures), the array-trained classifiers achieved an average F1 of 0.98 and the MethaDory-PacBio classifiers an average F1 of 0.96.

Similarly to our test set, the lower performances were concentrated in several signatures, e.g. one of the *CHD8* signatures^124^, or the moderate-effect size signatures for *ANKRD11*^148^ and *CHD3*^184^. These results are in line with the internal benchmarking and confirm the performance of the MethaDory classifiers also in external cohorts.

To implement MethaDory for clinical testing, we performed *in-house* MethaDory clinical validation. We enrolled in parallel a prospective, blinded validation cohort of 39 consecutive individuals submitted for the current standard-of-care array-based DNAm testing provided at EpiSign^TM^ V5 (Amsterdam UMC). MethaDory classifiers obtained a 100% concordance for these samples with EpiSign^TM^, classifying 5 as a case for 5 disorders (*CDK13*, *CHD4*, chr7q11.23dup, *BRWD3*, and *TET3*) and 34 as controls (F1=1, **Supplementary Table 18**).

Across internal and external validation, 154/188 episignatures for 82 disorders were tested on at least one affected individual. Among these, 134 episignatures achieved an F1>0.80 (101 with F1>0.95), while the remaining had lower performance. These results support the need for further validation of the untested episignatures and for the refinement of the underperforming ones.

### Clinical utility of MethaDory

#### MethaDory supports variant reclassification of variants of uncertain significance

Given that episignatures are currently mainly used to assist in reclassification of VUS, we assessed the agreement of MethaDory, EpigenCentral, and NSBEpi for clinically challenging variants from 95 samples carrying a VUS (**Supplementary Table 19, Figure 4A-B**). All three approaches had 97% concordance, with 63 being classified by all three as controls and 29 as cases. One *CHD8* sample (GSM3124840) was classified as affected case by the majority (2/3) of the available *CHD8* episignatures in MethaDory and EpigenCentral, but was predicted as control by the NSBEpi. This is because the Aref-Eshghi et al., published *CHD8* episignature performance does not replicate in NSBEpi, in MethaDory, and in previous independent study^119^. Similarly, only MethaDory and EpigenCentral classified *NSD1* sample (GSM1920465) as a case, in agreement with the original report^102^. Conversely, MethaDory together with NSBEpi classified a sample (GSM3572717) as *SMARCA2* case, but EpigenCentral had intermediate score. The individual had milder clinical phenotype than typically reported for NCBRS and carried the variant NM_003070.5(*SMARCA2*):c.3849G>T p.(Trp1283Cys)^95^, which was previously reported to present with atypical DNA methylation changes, affecting only some of the *SMARCA2*-associated CpGs^95^. Therefore, intermediate scores generated by the EpigenCentral and two of the *SMARCA2*-classifiers in MethaDory better reflect the atypical nature of this variant. These results highlight the utility of using multiple signatures for the same disease, derived from different cohorts and studies. Where signatures disagree, their derivation (sample size, variant representation) and independent performance should be weighed alongside molecular and clinical evidence^12,19,119^.

**Figure 4.**
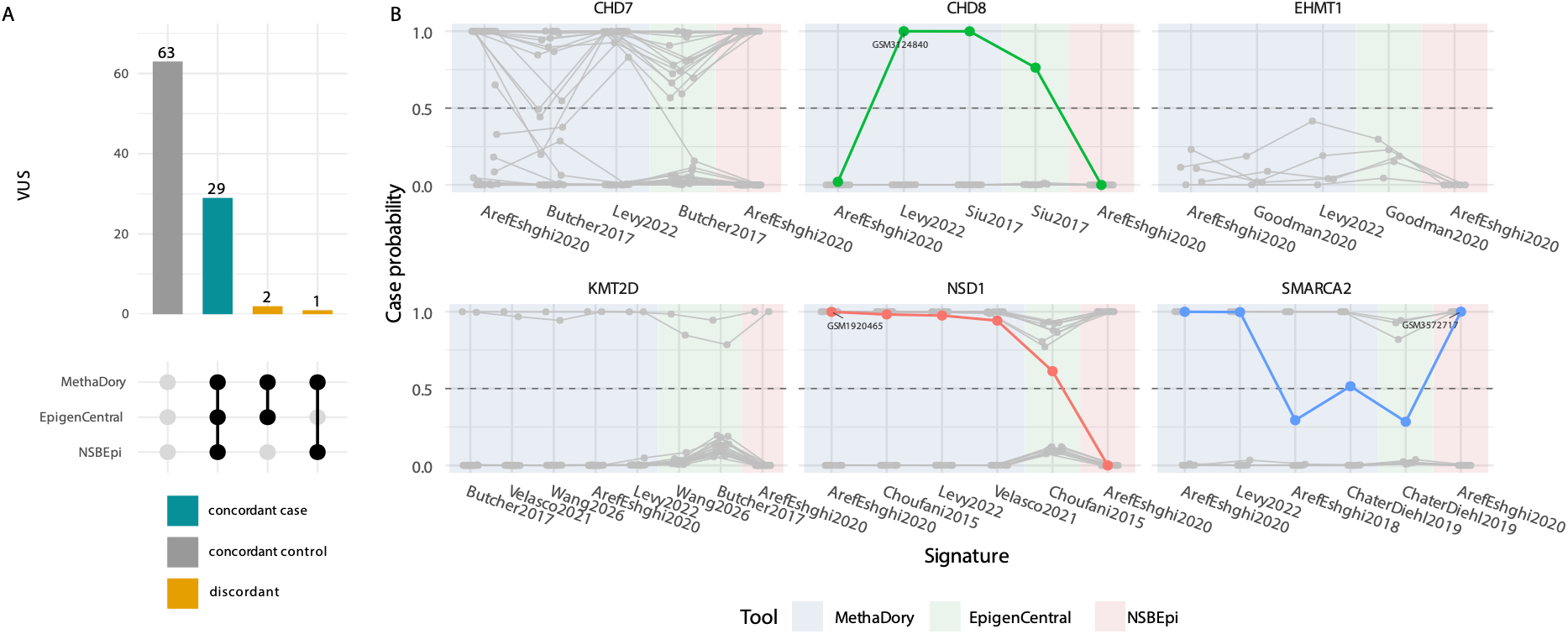
VUS reclassification for MethaDory, EpigenCentral and NSBepi. **A)** Upset plot showing the agreement in reclassification of VUS by MethaDory, EpigenCentral, and NSBepi. **B)** Prediction scores for the samples carrying VUS previously reclassified with DNAm analysis. Each dot represents the prediction score for a specific sample, over the available classifiers trained in this study with synthetic cases or present in EpigenCentral or NSBepi. Samples are stratified by gene. Since NSBepi does not generate a probability score, predictions of “control” were assigned a value of 0 and predictions of “disease” a value of 1.

#### MethaDory facilitates ultra-rapid diagnostics

We previously highlighted the utility of DNAm testing in ultra-rapid diagnostic settings, expediting variant curation and interpretation for a CHARGE syndrome patient^188^. MethaDory has been integrated in our diagnostic ONT ultra-rapid service and facilitated the diagnostic process in three additional cases after the original publication.

The first case was a newborn presenting with microcephaly, thick vermilion border, aortic, and pulmonary stenosis. MethaDory testing was positive for the Williams syndrome episignature, which was confirmed by identifying 7q11.23 deletion within 124 hours.

The second case was a newborn presenting with craniofacial dysmorphism, tall stature, abnormal genitalia, congenital heart disease, and a supernumerary nipple. Brain imaging revealed cerebral white matter hypoplasia and dysplasia of the superior cerebellar vermis. Trio ultra-rapid ONT genome sequencing returned in ∼100 hours, and revealed compound heterozygous pathogenic variants in *PMM2,* confirming a diagnosis of congenital disorder of glycosylation, type Ia. Additionally, *de novo ARID1B* variant was identified NM_001363725.2(*ARID1B*):c.4264A>G p.(Met1422Val). This variant is located outside of domains, was not predicted to affect protein structure or splicing (AlphaMissense = 0.053, SpliceAI = 0) MethaDory classified the sample as control, which together with the clinical presentation and the alternative diagnosis, supported that the *ARID1B* variant was likely benign and does not contribute to the patient’s phenotype.

The third case was a newborn presenting with multiple congenital anomalies, including congenital heart disease, butterfly and thoracic hemivertebrae, and epispadias. Ultra-rapid ONT genome sequencing and extensive reanalysis did not identify a causative variant. However, MethaDory returned a positive classification for the RCEM episignature, supporting that clinical diagnosis, in addition to an alternative molecular cause.

#### MethaDory helps solving unsolved individuals with rare disorders

MethaDory has also been included in the re-analysis pipeline that we designed for unsolved individuals tested at the Erasmus MC. We report three cases where MethaDory-aided episignature analysis together with long-read sequencing was used for the diagnosis.

We were referred two individuals presenting with a clinical phenotype of CHARGE syndrome, but negative molecular genetic testing. In the first case, EpiSign^TM^ assay returned a positive classification result for the *CHD7* episignature. Reanalysis of exome sequencing data remained inconclusive. We then performed ONT long-read genome sequencing. MethaDory confirmed the positive *CHD7* episignature result, and, further identified a deep intronic *CHD7* variant, NM_017780.4(*CHD7*):c.5210+1091T>G, p.(?), which is absent from gnomAD v4.1.1, and predicted to affect splicing by leading to the inclusion of a cryptic pseudoexon (SpliceAI-AG 0.17, SpliceAI-AL 0.13, Pangolin splice gain 0.37, **Supplementary Figure 15**). For the second case, ONT long-read genome sequencing revealed the variant NM_017780.4(*CHD7*):c.4533+937T>G, p.?, also absent in gnomAD v4.1.1., predicted to lead to the inclusion of an out-of-frame pseudoexon (SpliceAI- AG 0.82, SpliceAI-DG 0.89, Pangolin splice gain 0.79, **Supplementary Figure 15**). MethaDory testing returned positive results across all available *CHD7* episignatures, providing support for classification of the variant and confirming a diagnosis of CHARGE syndrome.

Finally in 2026, Erasmus MC held a *Solveathon* for undiagnosed patients with suspected rare monogenic disease. A total of 13 out of the 33 affected participants had DNAm data via DNAm arrays (n=5) or ONT (n=8). Among these, one case presented with esophageal atresia, hip dysplasia, congenital heart disease, splenomegaly, intellectual disability, intestinal pseudo-obstruction, hypo/aplastic thumbs, and, after negative genetic testing, MethaDory revealed “positive” RCEM episignature. Given the absence of other molecular causes, the clinical presentation and “positive” episignature results supported the diagnosis of VACTERL association.

## Discussion

In this study, we developed MethaDory, a framework to train rare disease episignature classifiers from synthetic epigenomes derived from controls alone, to classify both DNAm arrays and long-read methylation profiles. MethaDory was validated internally and externally (in 7 centres) and the classifiers performed on par with or outperformed the available alternatives, while greatly expanding the repertoire of openly available signatures to 102 disorders, compared with the <40 in the publicly available approaches, EpigenCentral^123^ and NSBEpi^21^. The companion MethaDory app runs completely locally, requiring no sharing of patient data, it can be easily expanded with more signatures as they are published, using the accompanying codebase, and can be expanded to additional technologies without requiring the (re-)sequencing of the affected individuals. The app also generates reports including heatmaps, and dimensionality-reduction plots, aiding the interpretation of the classifiers results as previously recommended^19^.

We further demonstrated the clinical utility of MethaDory episignature classifiers in rare disease research and diagnostics, including the simultaneous analysis of genetic variants (short variants and structural variants) and DNAm profiles from long-read data (both ONT and PacBio). MethaDory helped to solve previously unsolved cases by integrating episignatures with long-read genome variant identification, including a case identified through the RCEM episignature during the Erasmus MC Solveathon, and two cases where a positive *CHD7* episignature consolidated a diagnosis of CHARGE syndrome caused by deep intronic variants. We show how episignature testing within the ultra-rapid genome sequencing program for the critically ill, corroborated three diagnoses, including the downgrading of a VUS in a critically ill newborn. Together, these cases show that MethaDory supports variant interpretation, resolves diagnostic odysseys, and accelerates diagnoses, while providing functional classification not only from DNAm array, but directly from the long-read sequencing DNAm data, helping realize the single-test paradigm recently proposed for ‘near perfect genome sequencing’^189^.

The synthetic data strategy in MethaDory addresses multiple limitations that hinder the wide adoption of episignatures^19^. First, it mitigates data scarcity in valid training examples, which is a major constraint for ML models training^22^, by expanding training sets by orders of magnitude for any utilised technology beyond the limited number of affected-individual samples available from each (ultra)rare disorder. This enables episignatures to be ported to potentially any current or future platform, decoupling the episignature availability from the platform it was generated, smoothing the ongoing transition to novel technologies. Second, by sampling across age groups, ethnic backgrounds, and sexes, it might improve classifier generalizability. Third, episignatures are typically derived and trained, from only typically presenting affected individuals, which reduces sensitivity for hypomorphic and mosaic variants^118^. By incorporating effect-size variability into synthetic data generation, we show that classifier performance can be enhanced even relative to models trained on affected-individual data^123^. Fourth, synthetic cases preserve affected individuals’ privacy by preventing the need to share sensitive and personal data. Importantly, our approach enables simultaneous comparison and validation of multiple signatures generated by different approaches, strategies, and cohorts. For example, of the three signatures for *CHD8,* one^124^ failed to classify all the positive cases while the other two signatures classified them correctly. The same episignature achieved also a sensitivity of 0 in NSBEpi, and another study reported the low sensitivity of this signature and advised against its usage in clinical settings^119^. These comparisons suggest that the quality of the underlying episignature determines the performance of episignature classifiers irrespectively of the training strategy. We also observed that some published episignatures, e.g., *CUL3*, *KDM4B*, and *PURA* failed to work with our approach and that the performance correlated with the episignature discriminatory power. We noted that some of these signatures had failed the leave-one-out cross validation in the original study (e.g, *CUL3*)^171^, or were derived using control samples assayed in batches separately from the cases (*PURA*)^164^, suggesting that technical confounders might affect the reported sites and effect sizes. Finally, the *KDM4B* episignature was also withdrawn from the commercial testing by EpiSign^TM^ V6^190^, as insufficiently robust for diagnostic use (private communication). This is in line with previous evidence supporting that not all signatures perform equally well^119^ and require independent validation before clinical application.

Importantly, episignature loci and classifiers are product of the affected individuals and methods used to derive them, and not the disease biology alone^191^. For the same reason, variant interpretation using DNAm signatures is not binary^19^ and intermediate scores can reflect mosaicism, hypomorphic, domain-specific variants, and milder phenotypes^19,191^. Conversely, a control classification in an individual carrying a (likely) pathogenic variant should be evaluated with the variant itself and the phenotype^9^. Moreover, episignatures are known to overlap for related genes, like components of protein complexes^19^, and are derived without accounting for other disorders, which may result in spurious classification results^12^. In line with this, Smits et al. reported a positive EpiSign^TM^ result in 9.0% (20/223) of DNA-testing unsolved individuals assessed in routine diagnostic setting^9^. However, only 11/20 individuals had matching clinical presentation and in 9/11, a causative variant or an imprinting defect could be retrospectively confirmed in the episignature-reported gene/locus. Smits et al. reported also that EpiSign^TM^ detected ∼90% of the cases with LP/P variants in the episignature associated genes, similarly to what we report for MethaDory for the in-house and independent multi-centre cross-platform validation. This supports that while the results of the episignature testing can increase diagnostic yield, its results should be interpreted together with other clinical and variant data^12,19^.

This work also gives an opportunity to reflect on how the field should share its outputs to ensure reproducible results and clinical validation. As the described disorders with an episignature are individually (ultra)rare, assembling cohorts large enough to derive and validate robust DNAm classifiers requires international efforts over years. Ideally, these efforts should result in data that are shared in an open and FAIR (Findable, Accessible, Interoperable, Reusable) manner. The tightened privacy regulations and the closed commercial entities with competing interests make this challenging, even though affected individuals themselves generally view data sharing positively, particularly for not-for-profit research^192^. While the individual-level data require strict access policy based on local and international regulations, we argue that the analytical results (episignatures loci with their effect sizes and confidence metrics, the trained classifiers and underlying code) should be openly shared for independent validation. This will also ensure that signatures can be potentially combined across cohorts and laboratories and reproducibly used to their full utility across the technologies for the rare disease community. Our external validation illustrates this model, by sharing the trained classifiers rather than patient data, seven centers could independently evaluate the classifiers on their own cohorts, while all individual-level data remained within the generating institutions. As an alternative that protects identity while enabling open sharing, we propose releasing a genome-wide cohort “DNAm metaprofile”, i.e. average profiles of cases and controls, and/or the effect size, including a per site variability metric, inspired by current standards for genome-wide association studies^193^. This would protect the identity of the affected individuals, while enabling their open-source sharing ^191^ and validation via approaches like MethaDory.

A limitation of our study is that it relies on publicly available episignatures whose derivation and reporting practices are not standardized: most studies report only statistically significant differentially methylated probes, with inconsistently calculated effect sizes and p-values, often further filtered for the classification task based on study-specific criteria. These prefilterings obscures disorder biology, preventing comparisons across disorders, and can degrade performance on external data. Additionally, as the reported sites are selected per disorder, the methylation of affected individuals at the sites of other signatures is unknown, which prevents the generation of synthetic cases of other disorders to train one-versus-all classifiers, as done with real affected individuals and that might help reduce episignature cross-reactivity. Some studies do not provide even this minimal information (even on personal request), preventing external validation. Despite the rarity of the disorders associated with episignatures, we obtained positive controls for most of the classifiers (154/188). Additional validation of the untested episignatures is required before they can be implemented into routine diagnostics and further development and refinement for the episignatures with lower performance, as this and previous studies have shown that published episignatures may not always reproduce reliably^119^. Finally, we observed that long-read synthetic epigenome MethaDory classifiers have a lower performance compared to the array-derived ones. We note that the amount of currently available long-read control epigenomes is far less diverse and limited compared to what is available for arrays but expected to grow rapidly with the clinical adoption of long-read sequencing. Additionally, we observed worse performance on the PacBio data than with ONT which may reflect the smaller PacBio test set, and the lower agreement of PacBio methylation calls with arrays than ONT calls (Pearson r 0.91 vs 0.98), in line with previous data^117,194,195^.

In summary, we demonstrate that synthetic epigenomes can train high-performance, cross-technology episignature classifiers overcoming the sample size, diversity, and data access limitations of the current strategies. We release these classifiers and MethaDory app openly for public use, extension, and validation. We also showcase clinical utility of MethaDory integrating episignature detection with long-reads genome sequencing to solve unsolved patients and facilitate ultrarapid diagnostics in a single test.

## Data and Code Availability

MethaDory app is available in GitHub (https://github.com/f-ferraro/MethaDory). Publicly available studies used in this work can be found in GEO using the identifiers listed in Supplementary Table 1, pre-processed data is available at https://zenodo.org/records/23033263 Trained classifiers are available in Zenodo https://zenodo.org/records/23038390. Episignature loci and effect sizes collected from the literature are also made available as UCSC Genome Browser track https://api.genome-euro.ucsc.edu/cgi-bin/hgTrackUi?g=methaDory.

Codebase for the analyses described will be available in GitHub upon publication and is attached a supplementary file for revision. Internal data and raw sequencing datasets are not available according to privacy regulations.

## Ethics statement

Research was conducted in line with the Declaration of Helsinki and approved under institutional board MEC-2025-0094 and MEC-2025-0194.

## Epi2Diag collaborators

Alexandra Afenjar, Jean-Luc Alessandri, Chloé Angelini, Thibaud Armand, Stéphanie Arpin, Benoit Arveiler, Séverine Bacrot, Catherine Badens, Sarah Baer, Claire Bar, Mouna Barat, Giulia Barcia, Magalie Barth, Nathalie Bednarek, Frédéric Bilan, Sahra Bodo, Céline Bonnet, Marie-Noëlle Bonnet Dupeyron, Elise Boucher, Franck Bourdeaut, Lucile Boutaud, Odile Boute, Anne-Claire Brehin, Coralie Briand, Céline Bris, Sophie Brisset, Adelaïde Brosseau-Beauvir, Tiffany Busa, Kévin Cassinari, Roseline Caumes, Lucile Cejudo, Camille Cenni, Pascal Chambon, Perrine Charles, Nicolas Chassaing, Nicolas Chatron, Nicole Chemaly, Bertrand Chesneau, Benjamin Cogne, Cindy Colson, Valérie Cormier Daire, Auriane Cospain, Nathalie Couque, Juliette Coursimault, Thomas Courtin, Charles Coutton, Isabelle Creveaux, Benjamin Dauriat, Maud De Dieuleveult, Wallid Deb, Geoffroy Delplancq, Bénédicte Demeer, Florence Demurger, Anne-Sophie Denomme-Pichon, Christel Depienne, Anne Dieux, Martine Doco-Fenzy, Véronique Duboc, Christèle Dubourg, Yannis Duffourd, Morten Dunoe, Romain Duquet, Benjamin Durand, Salima El Chehadeh, Camille Engel, Lucas Fares-Taie, Patricia Fergelot, Mélanie Fradin, Christine Francannet, Livia Garavelli, Aurore Garde, David Genevieve, Anna-Gabriella Gerasimenko, Emmanuelle Ginglinger, Cyril Goizet, Alice Goldenberg, Mihelaiti Guberto, Paul Gueguen, Anne-Marie Guerrot, Anne Guimier, Trine Bjoerg Hammer, Abdool Majid Hasan Miyan, Solveig Heide Guihard, Lucas Herissant, Delphine Heron, Clara Houdayer, Céline Huber, Bertrand Isidor, Aurélia Jacquette, Clémence Jacquin, Louis Januel, Guillaume Jouret, Sophie Julia, Boris Keren, Paul Kuentz, Marilyn Lackmy, Didier Lacombe, Fanny Laffargue, Laetitia Lambert, Alinoe Lavillaureix, Pauline Le Tanno, François Lecoquierre, Marine Legendre, Bruno Lehup, Gaëtan Lesca, Marion Lesieur-Sebellin, James Lespinasse, Jonathan Levy, Laurence Lode, Valérie Malan, Caterina Marconi, Henri Margot, Valentine Marquet, Pauline Marzin, Isabelle Maystadt, Lara Mellinger, Sandra Mercier, Vincent Michaud, Cyril Mignot, Mathieu Milh, Marie-Pierre Moizard, Pauline Monin, Godeliève Morel, Jérémie Morteux, Sophie Nambot, Sophie Naudion, Caroline Nava, Gaël Nicolas, Mathilde Nizon, Antonio Novelli, Sylvie Odent, Laurence Olivier-Faivre, Julien Osouf, Mathilde Pacault, Philippe Parent, Ilaria Parenti, Laurent Pasquier, Olivier Patat, Alix Paulet, Cristina Peduto, Julie Pernin-Grandjean, Florence Petit, Nicole Philip, Juliette Piard, Leticia Diana Pias Peleteiro, Jean-Marc Pinard, Elide Pisan, Amélie Piton, Marc Planes, Pauline Plante-Bordeneuve, Céline Poirsier, Linda Pons, Audrey Putoux, Chloé Quelin, Sylvia Quemener, Caroline Racine, Mélanie Rama, Francis Ramond, Laure Raymond, Mathilde Renaud, Lucile Riera-Navarro, Marlène Rio, Thomas Rio Frio, Paul Rollier, Sophie Rondeau, Caroline Rooryck, Massimiliano Rossi, Joëlle Roume, Flavien Rouxel, Valentin Ruault, Hana Safraou, Sofia Said, Maj-Britt Salewski, Maria José Sanchez Soler, Pascale Saugier-Veber, Elise Schaefer, Caroline Schluth-Bolard, Thomas Smol, Myriam Soleimani, Brian Sperelakis Beedham, Radka Stoeva, Mylène Tharreau, Christel Thauvin, Renaud Touraine, Annick Toutain, Frédéric Tran Mau Them, Christine Tranchant, Kévin Uguen, Julien Van Gils, Roxane Van Heurck, Clémence Vanlerberghe, Gabriella Vera, Alain Verloes, Catheline Vilain, Nathalie Villeneuve, Marie Vincent, Catherine Vincent-Delorme, Eléonore Viera-Dupont, Antonio Vitobello, Marie-Laure Vuillaume-Winter, Marjolaine Willems, Alison Yeung, Khaoula Zaafrane-Khachnaoui, Alban Ziegler

## Data Availability

MethaDory app is available in GitHub (https://github.com/f-ferraro/MethaDory). Publicly available studies used in this work can be found in GEO using the identifiers listed in Supplementary Table 1, pre-processed data is available at https://zenodo.org/records/23033263 Trained classifiers are available in Zenodo https://zenodo.org/records/23038390. Episignature loci and effect sizes collected from the literature are also made available as UCSC Genome Browser track https://api.genome-euro.ucsc.edu/cgi-bin/hgTrackUi?g=methaDory. Codebase for the analyses described will be available in GitHub upon publication and is attached a supplementary file for revision. Internal data and raw sequencing datasets are not available according to privacy regulations.

## Acknowledgements

We thank the affected individuals and their families that have contributed samples to all the studies we accessed, the many authors who contributed open-access data in GEO, and all authors that have also included episignatures and effect sizes of the episignature they published. We also want to thank Dr Mariëlle Alders, Amsterdam UMC, for providing DNAm raw data of individuals from Erasmus MC tested with EpiSign^TM^ at Amsterdam UMC.

The authors would also like to thank Dr Maximilian Haeussler for the inclusion of the episignature loci collected from the literature in the UCSC Genome Browser track.

## Funding

This research was supported by funding from the Netherlands Organisation for Health Research and Development (ZonMw) (TK, 91718310), Stichting Vrienden van het Sophia (EB & BMdG WAR23-37, DS & VJMV WAR24-34, JMAV & WTR WAR25-52), the Region Hovedstadens Forskningsfond til Sundhedsforskning (MH 2024-0059), and the National Institute for Health and Care Research (NIHR)

Manchester Biomedical Research Centre (NIHR203308). S.B. acknowledges the support of the MRC Epigenomics of Rare Diseases (EpiGenRare) Node (MR/Y008170/1). This work was supported from the project MULTIOMICS_CZ (Program Johannes Amos Comenius, Ministry of Education, Youth and Sports of the Czech Republic, ID Project CZ.02.01.01/00/23_020/0008540) - Co-funded by the European Union. LN was funded by the grant NU23-07-00281 and NW24-04-00067 from the Ministry of Health of the Czech Republic and thanks the National Center for Medical Genomics (LM2023067). M.Z.’s research is supported by a “Schlüsselprojekt” grant from the Else Kröner-Fresenius-Stiftung (2022_EKSE.185) and by a research grant from the German Research Foundation (DFG), project number 595395689 (ZE 1213/6-1). In addition, M.Z.’s omics research has received funding from the Federal Ministry of Education and Research (BMBF) and the Free State of Bavaria under the Excellence Strategy of the Federal Government and the Länder, as well as by the Technical University of Munich - Institute for Advanced Study. HP acknowledges the support by funding from the BMBF (German Federal Ministry of Education and Research) through the EJP RD project GENOMIT (01GM2301) - Co-funded by the European Union, and the German Center for Child and Adolescent Health (DZKJ) under the funding code 01GL2406B.

## Author contributions

Conceptualization: DR; Methodology: FF, DR; Software: FF; Investigation: FF, LB, MD, HCvdL, MdG, FP, AS, HR, MH, RS, IGvdH, NW, IG, MJ, IM, MRe, MRo, GT; Data curation: FF, LB, MD, HCvdL, MdG, FP, AS, HR, MH, ES, AM, DS, BMdG, RS, EB, AB, IGvdH, NW, IG, MJ, IM, MRe, MRo, GT, ZT, JV, CCh, GS, HP, FK, CG, TJvH, TK; Resources: SC, NC, EB, YvB, AB, LDK, MOM, NvE, AdB, TSB, WtR, JMAV, VJMV, NB, IG, MJ, IM, MRe, MRo, GT, LN, CCa, DG, IK, AW, MZ, MW, SB, RW, ZT, JV, CCh, GS, HP, FK, CG, TJvH, TK; Supervision: TJvH, TK; Project administration: DR; Funding acquisition: MH, DS, EB, AdB, JMAV, WtR, MZ, VJMV, LN, SB, HP, TK; Visualization: FF, DR; Writing – original draft: FF; Writing – review & editing: LB, MD, HCvdL, MdG, FP, AS, HR, MH, ES, SC, NC, AM, DS, PK, BMdG, RS, EB, YvB, AB, LDK, IGvdH, MOM, NvE, AdB, NW, RD, TSB, WtR, JMAV, VJMV, NB, IG, MJ, IM, MRe, MRo, GT, LN, CCa, DG, IK, AW, MZ, MW, SB, RW, ZT, JV, CCh, GS, HP, FK, CG, TJvH, TK, DR. All authors have read and approved the final manuscript.

## Author disclosures

MD, DR has received travel support from Oxford Nanopore Technologies.

Other authors have no conflicts of interest to disclose.

## LLM usage disclosure

Claude (Anthropic, Opus v.5.0-5.5) was used to polish the text language, linting, and annotating code. LLMs were not used to generate or modify data nor to make analytic or take interpretative decisions. All LLM-assisted content was reviewed, verified, and edited by the authors.

## Supplementary Figures

**Supplementary Figure 1.**
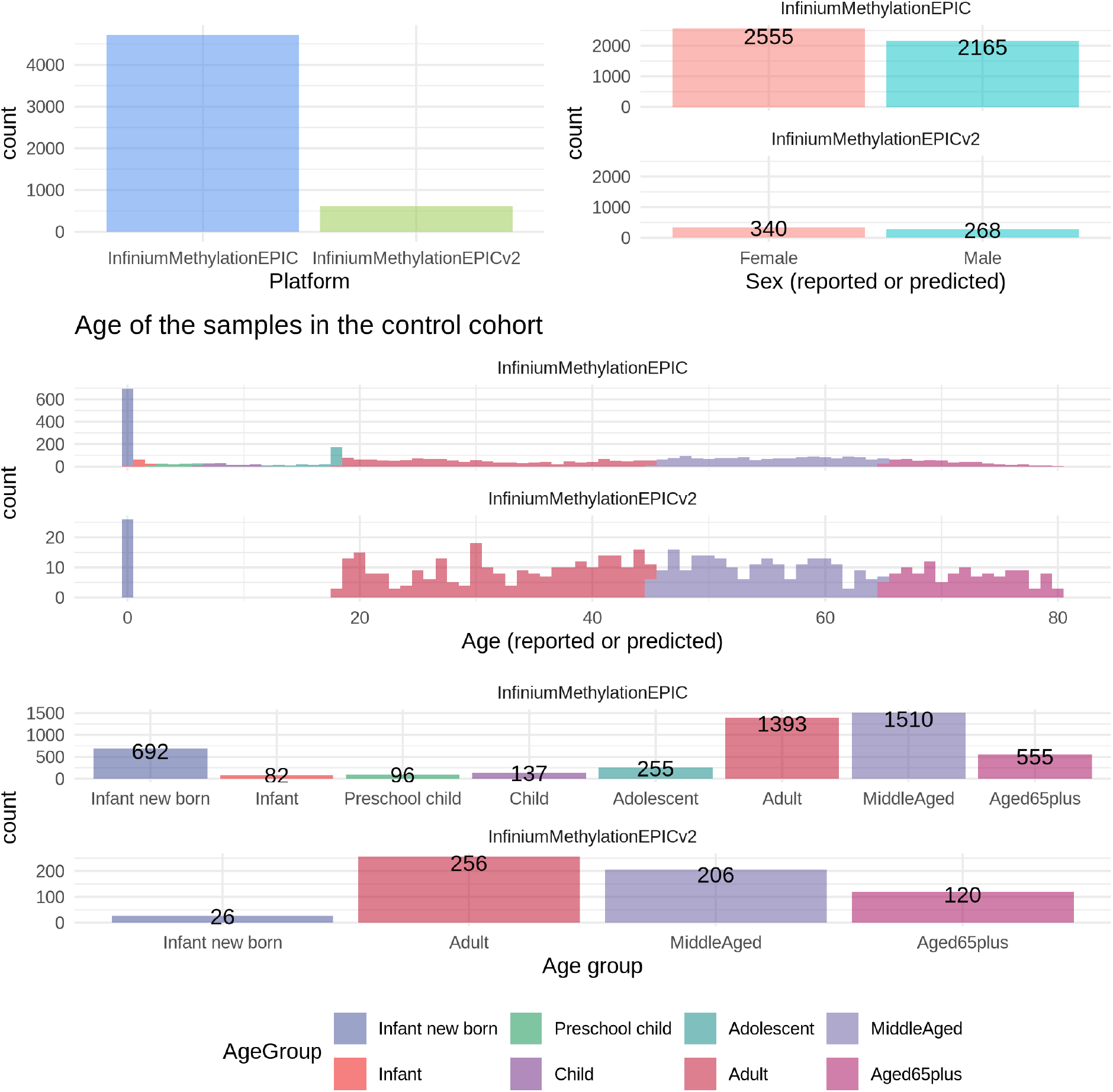
Demographics and characteristics of the control DNAm samples obtained from literature after quality control.

**Supplementary Figure 2.**
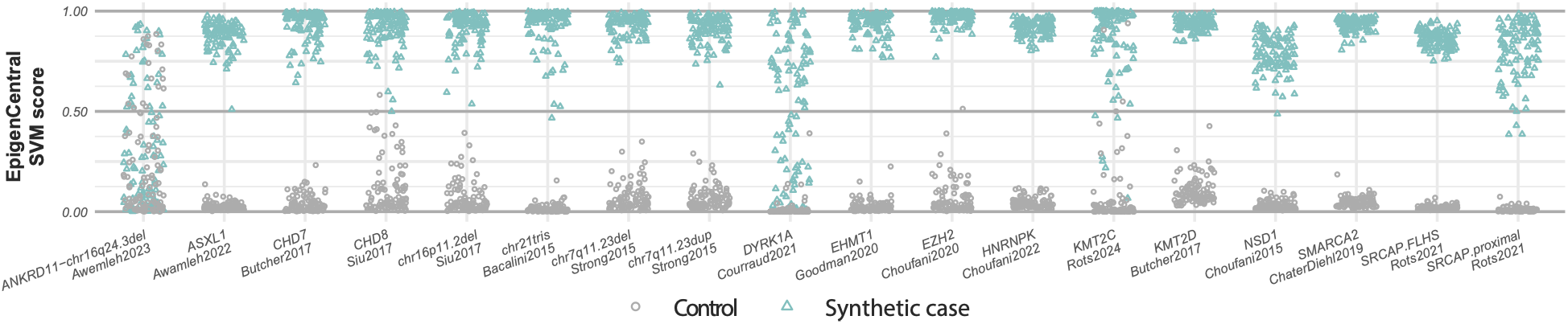
Synthetic cases are correctly classified by corresponding DNAm classifier present in EpigenCentral. Predictions of the DNAm classifiers for 18 diseases present in EpigenCentral for 100 unaffected individuals and 1,800 synthetic cases generated with the corresponding EpigenCentral DNAm signature. Each point represents a sample: empty circles represent an unaffected individual, and a triangle represents a synthetic case of the corresponding episignature. SVM score represents the probability of a sample being classified as a positive case for the specific DNAm signature.

**Supplementary Figure 3.**
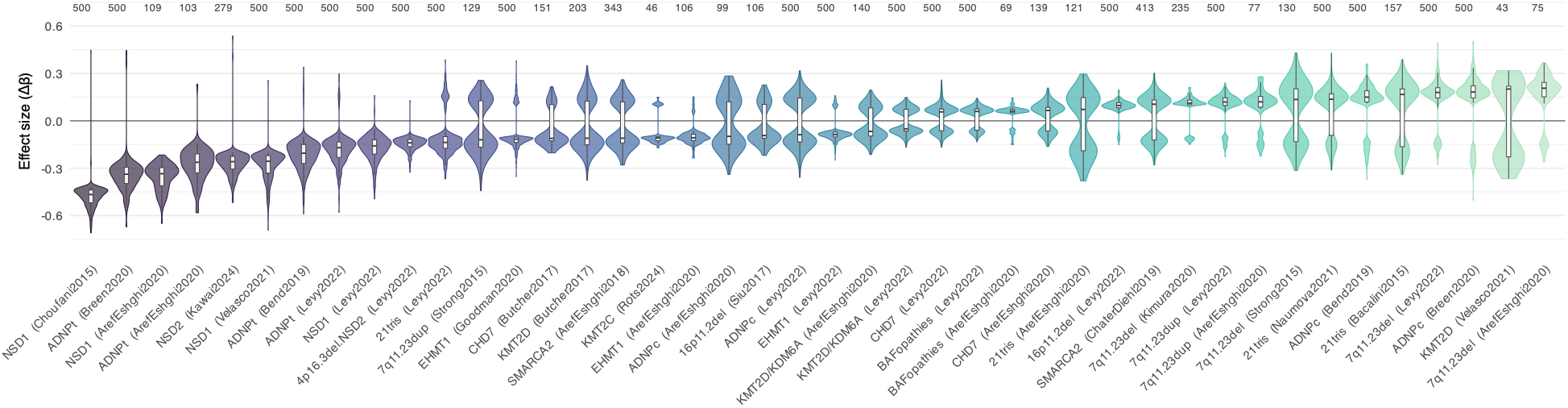
Reported effect size (delta beta) distribution of the 42 DNAm signatures used to optimize the synthetic data generation and classifier training strategy. The number of CpGs included in the signature is reported on top of each signature. Signatures are sorted by average effect size. This subset of disorders was representative of the breadth of reported DNAm signatures, with variable effect sizes and extent of effects.

**Supplementary Figure 4.**
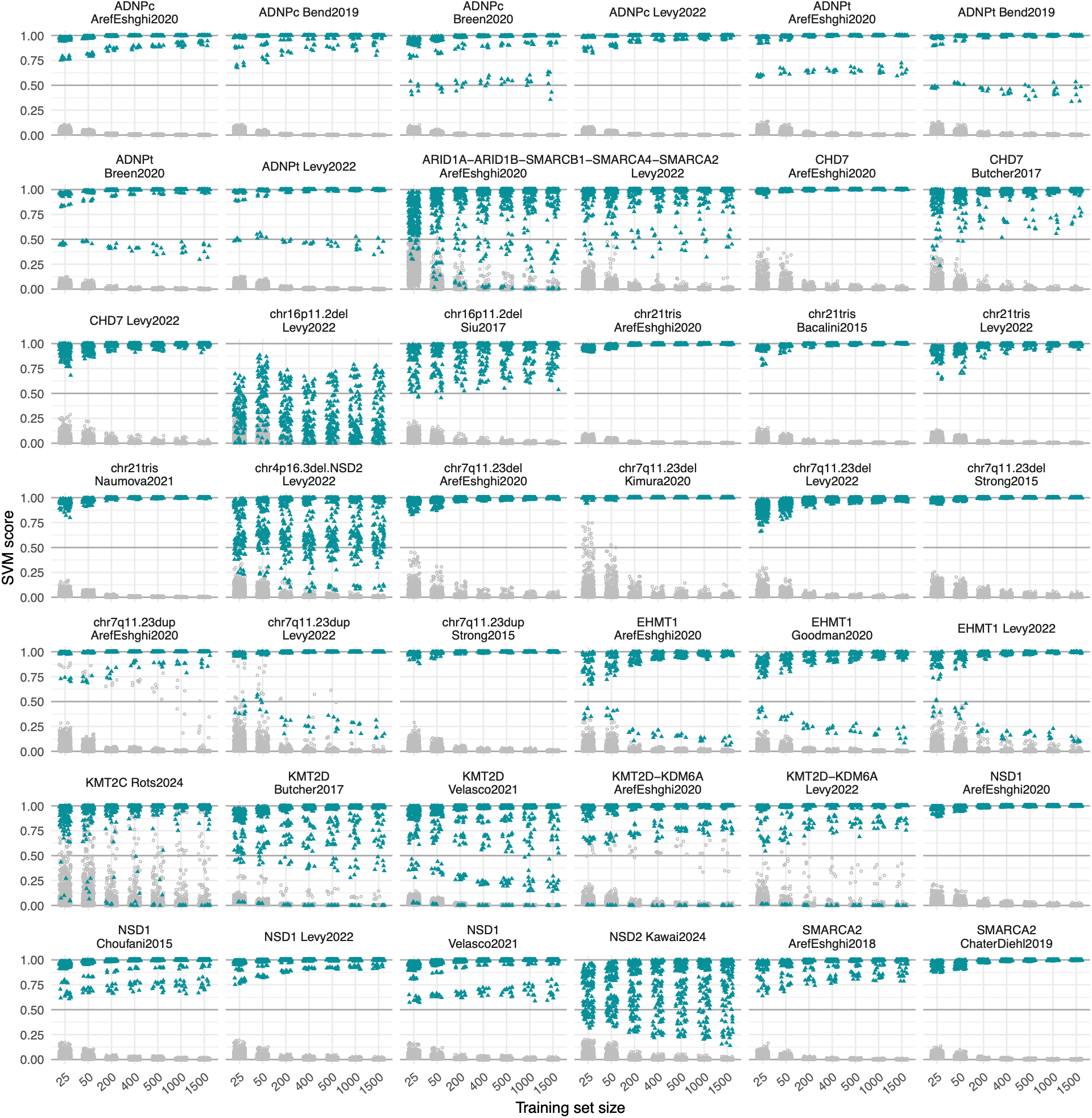
Impact of the size of synthetic training set on the performance of DNAm classifiers: Prediction results of 42 DNAm SVM classifiers trained with different training set sizes (x-axis) for predicting 213 affected individuals −17 ADNP syndrome (8 ADNPc, i.e. samples with variants within the central domain c.2000-2340, and 9 ADNPt, i.e. with variants outside c.2000-2340), 9 ARID1B syndrome, 17 CHD7 syndrome, 11 with a deletion of chr16p11.2 syndrome, 12 with chr21 trisomy, 38 with a deletion of chr7q11.23 syndrome, 7 with a duplication of chr7q11.23 syndrome,7 EHMT1 syndrome, 14 KMT2C syndrome, 20 KMT2D syndrome, 31 NSD1 syndrome, 16 NSD2 syndrome, 10 SMARCA2 syndrome, 1 SMARCA4 syndrome, 3 SMARCB1 syndrome - and 164 unaffected individuals. SVM score represents the probability of a sample being classified as a positive case for the specific DNAm signature. Affected individuals are shown in teal, controls in grey.

**Supplementary Figure 5.**
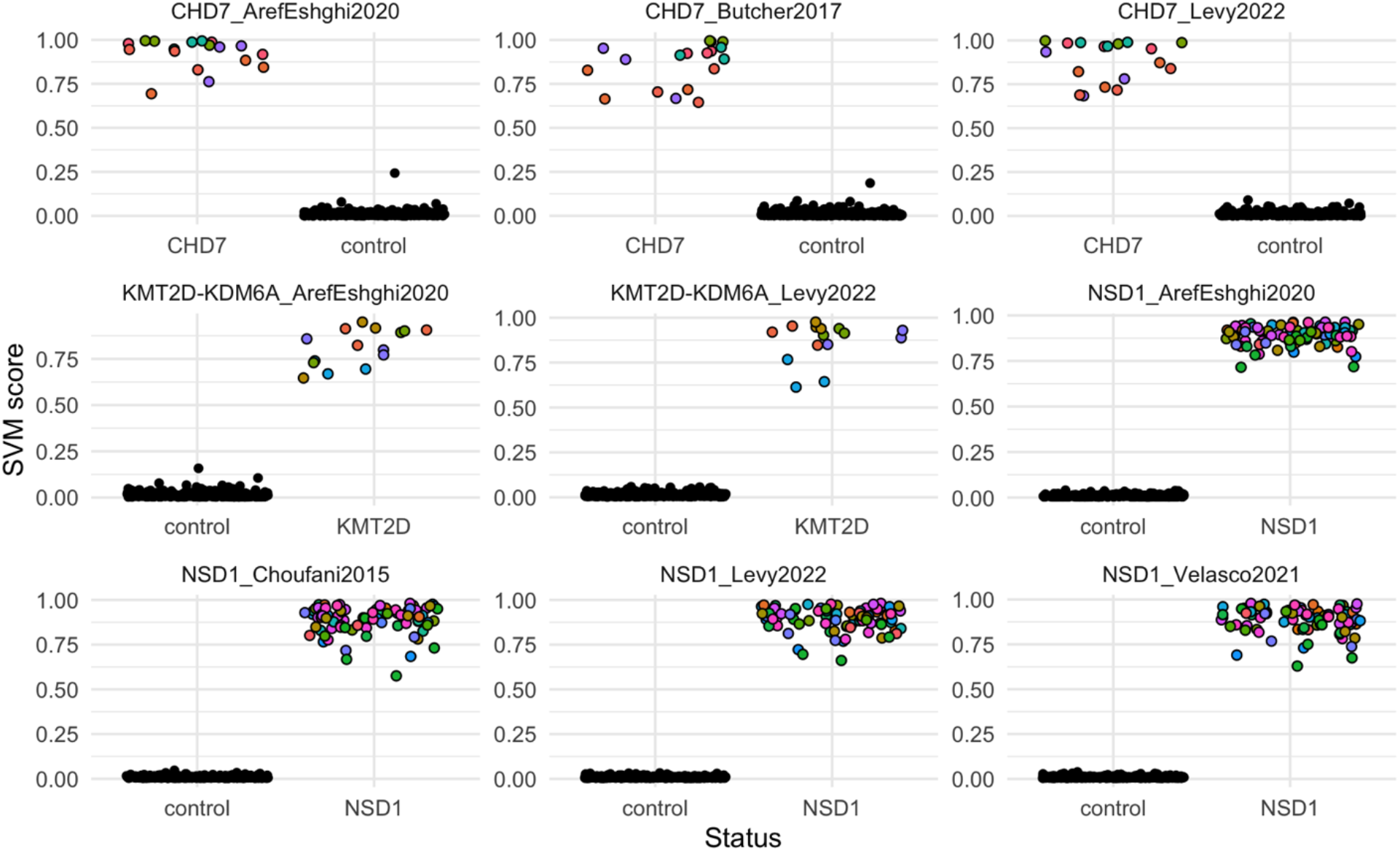
Variability of SVM scores for individual samples across independently trained classifiers. For each of three disorders (CHD7, KMT2D, NSD1), three SVMs were trained on the same DNAm signature using three non-overlapping subsets of 5 affected individuals, each combined with a non-overlapping subset of 100 controls (sampling without replacement). Each dot represents the SVM score obtained by an affected individual sample (indicated by different color within each signature) or a healthy control (colored in black) predicted by an SVM trained with different subsets of affected individual samples and controls but on the same DNAm signatures.

**Supplementary Figure 6.**
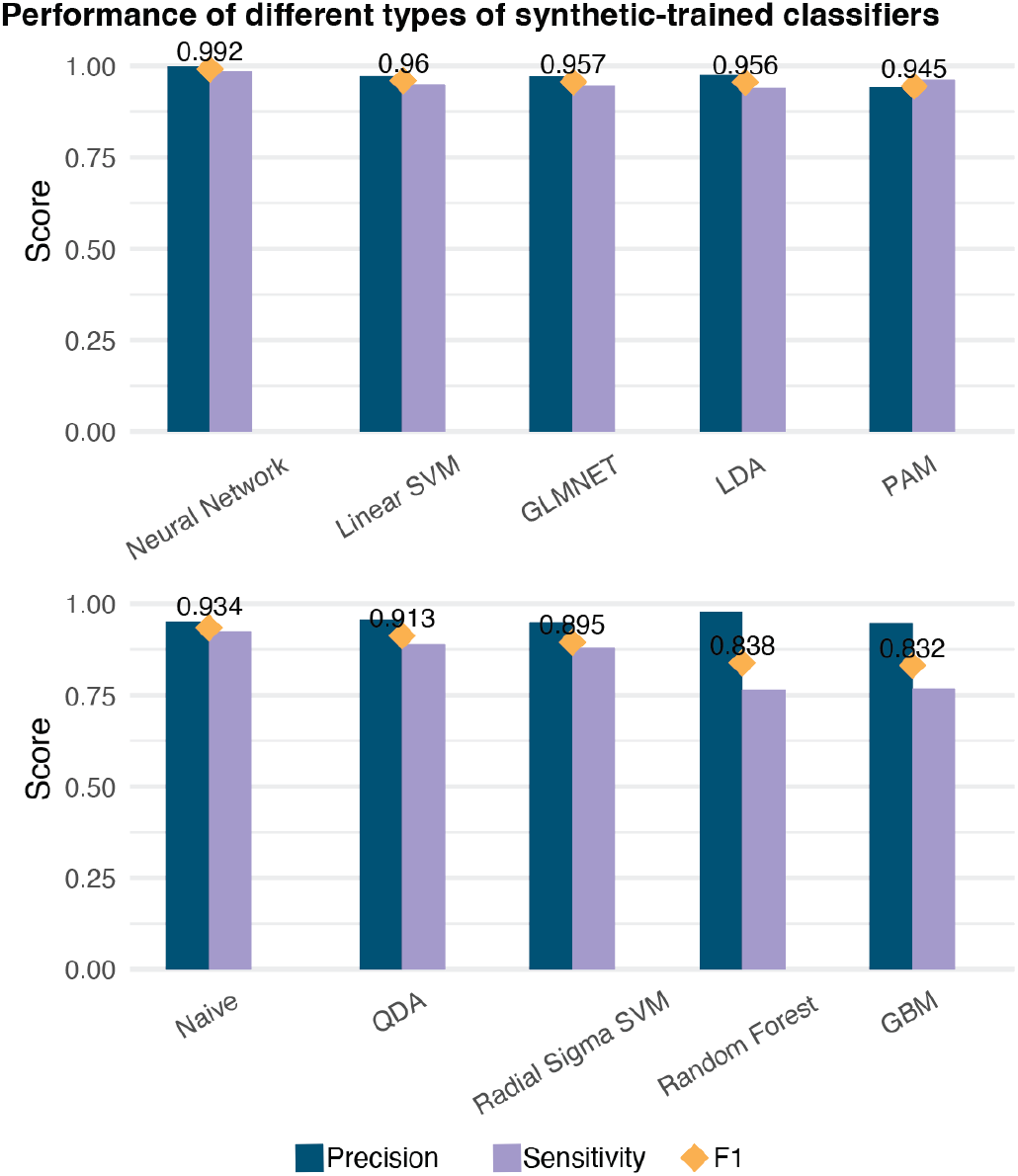
Comparison of machine-learning models trained on synthetic cases. Classification performance of the 10 models trained on synthetic cases and evaluated on affected individuals and controls. Since caret methods ‘svmLinear1’ and ‘svmLinear2’ converged on the same predictions for each sample, we only plotted a single summary one.

**Supplementary Figure 7.**
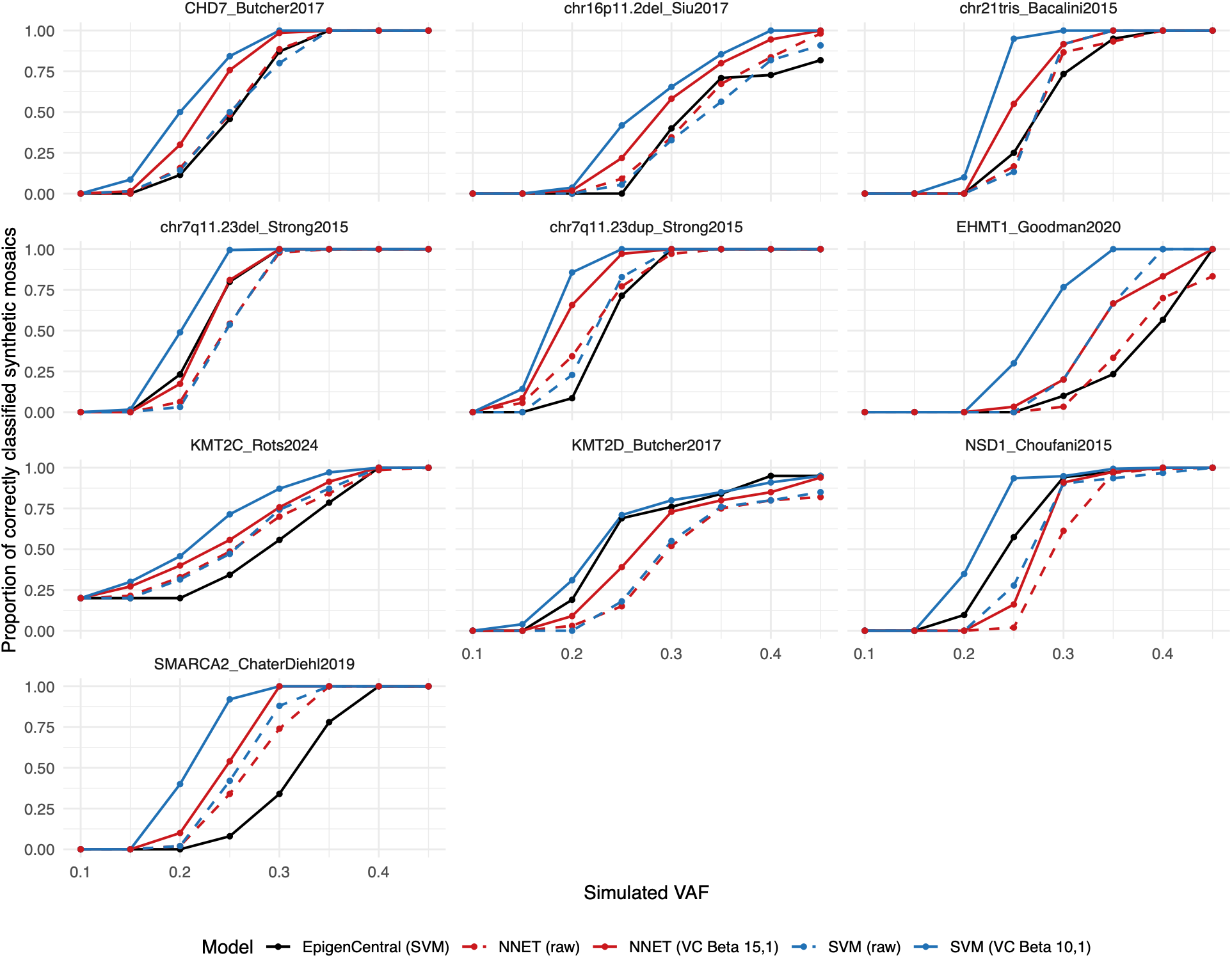
Variability coefficient modelling improves DNAm classifiers performance on intermediate effect variants and mosaics. Proportion of correctly classified synthetic mosaics at different simulated effect sizes, by MethaDory DNAm classifiers with different VC and by corresponding EpigenCentral DNAm classifiers. Results are reported for the models without the variability coefficient (VC), and for the best performing distributions.

**Supplementary Figure 8.**
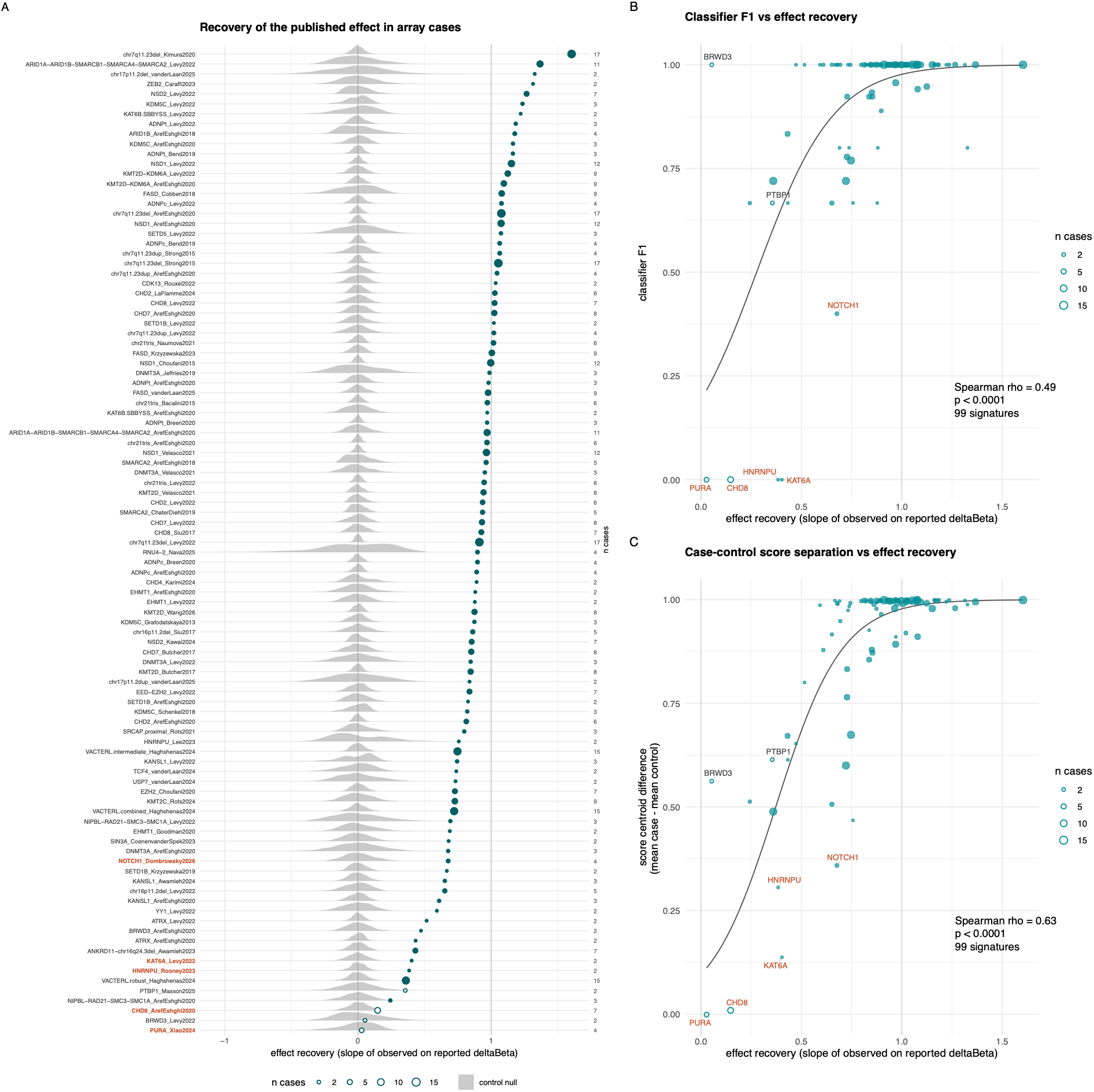
Recovery of the published episignature effects in affected individuals and its relationship with the performance of MethaDory-array classifiers. **A)** Effect recovery for each signature in the DNAm array test set. Each dot represents a signature, sized by the number of affected individuals, reported on the right axis. Grey halfviolins show the control null distribution, obtained by comparing randomly drawn sets of test controls with the remaining control over 1,000 resamplings. Hollow dots indicate episignatures with Pearson’s correlation between observed and published effects below the control null. Episignature whose classifier achieved F1 < 0.5 are shown in red. Relationship between effect recovery and **B)** the F1 score of the corresponding classifier and **C)** the difference between the mean classifier scores of affected individuals and controls. Each dot represents a signature.

**Supplementary Figure 9.**
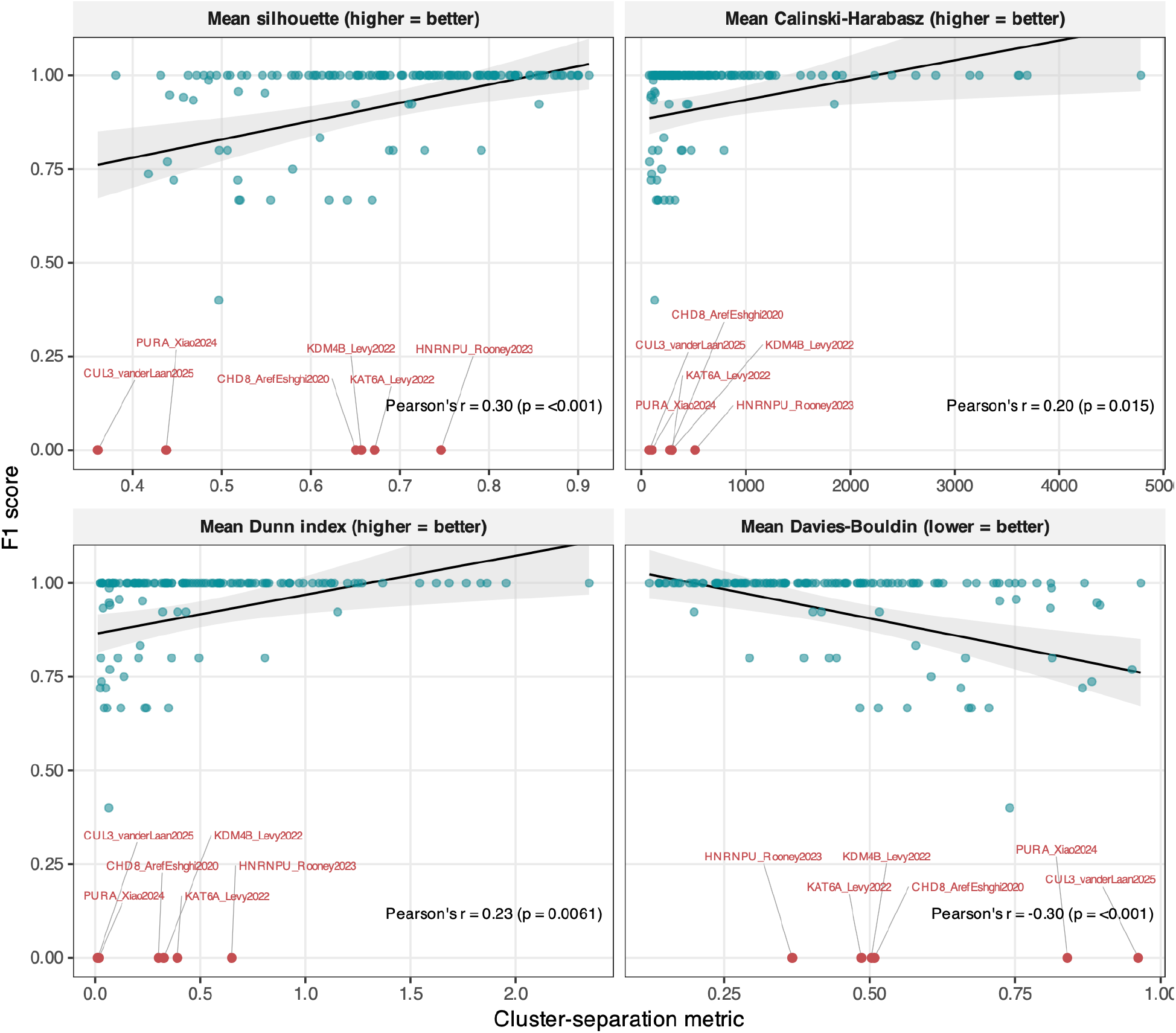
Relationship between cluster separation metrics (silhouette width, Calinski-Harabasz index, Davies-Bouldin index, and Dunn index) and the achieved F1 score of MethaDory-array classifiers on samples from affected individuals and controls. Each dot represents a signature. Signatures showing high silhouette width, Calinski-Harabasz and Dunn indices, but low Davies-Bouldin index are estimated to have strong discriminative capacity between simulated disease and control methylation profiles.

**Supplementary Figure 10.**
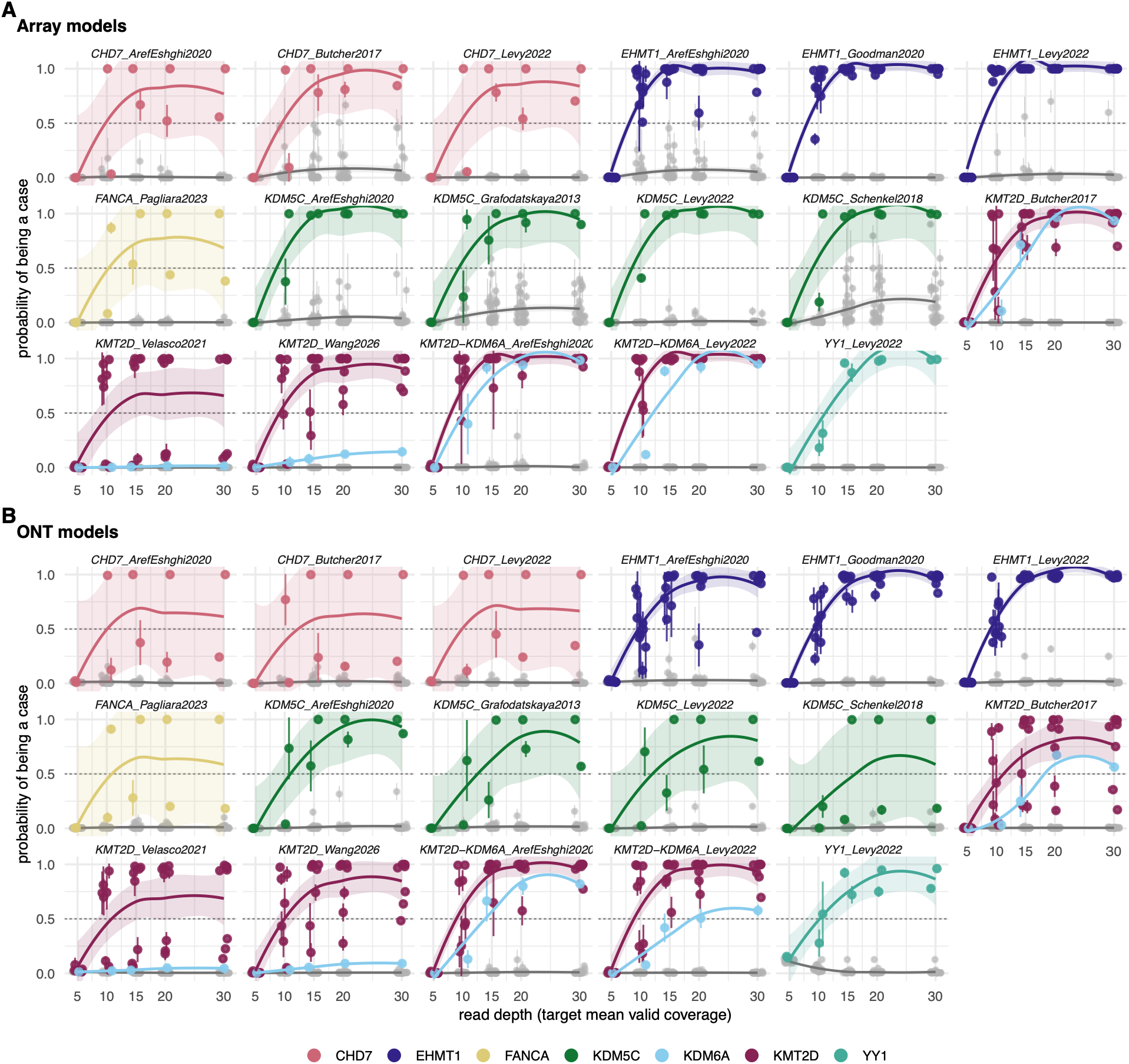
Effect of ONT read depth on MethaDory classification scores. Held-out ONT test samples (29 affected individuals and 28 controls) were downsampled to five target coverages three times. Points are individual samples, positioned at the median score across the three samplingsLines are loess trends with 95% confidence bands fitted per class. A) Models trained on DNAm arrays. B) Models trained on ONT.

**Supplementary Figure 11.**
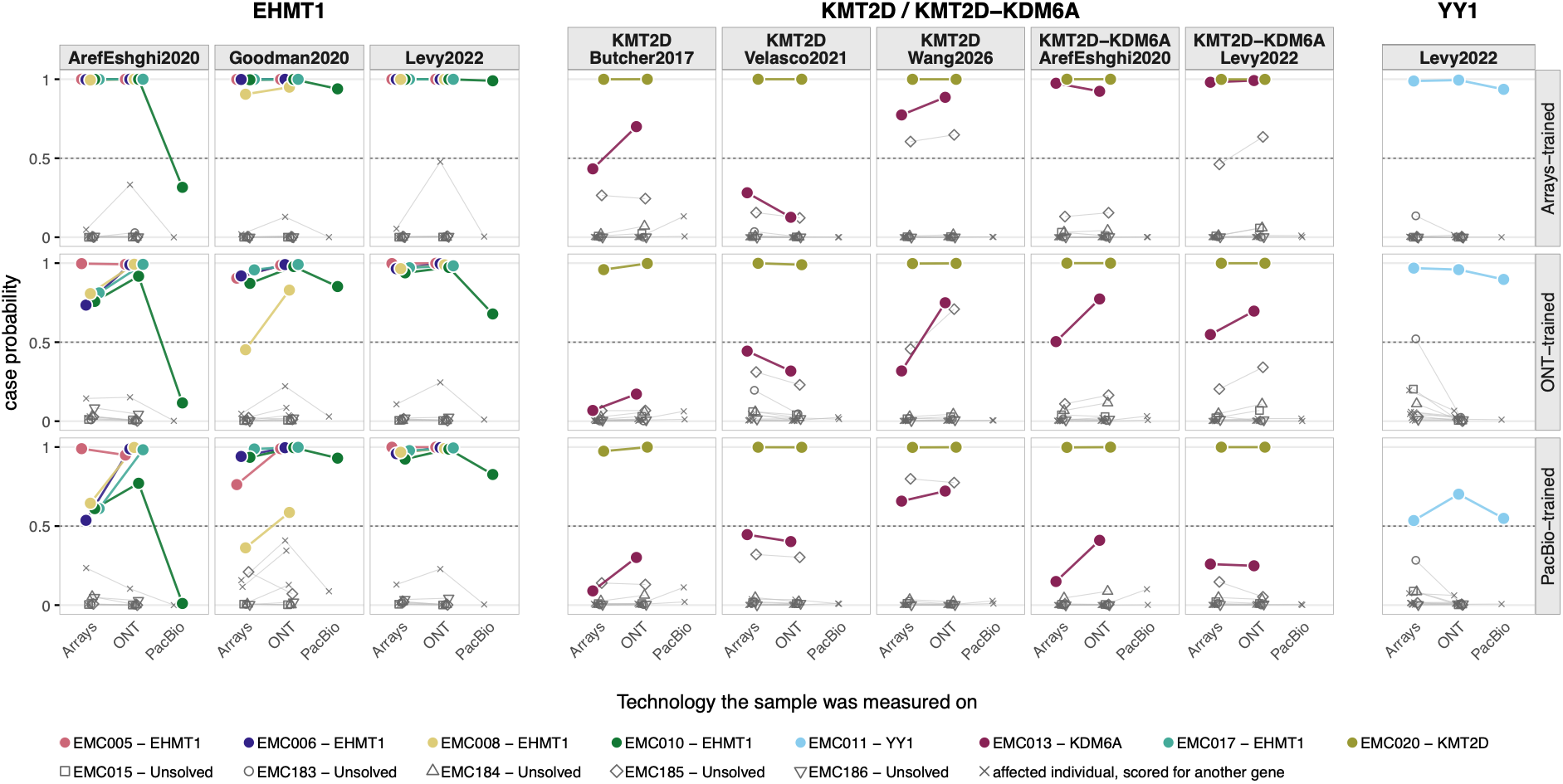
Prediction results for samples assessed with multiple platforms on models trained with synthetic cases from different technologies. Same DNA samples are connected with a line and color coded. For each disease, samples only corresponding affected individuals are color coded and other samples are in gray.

**Supplementary Figure 12.**
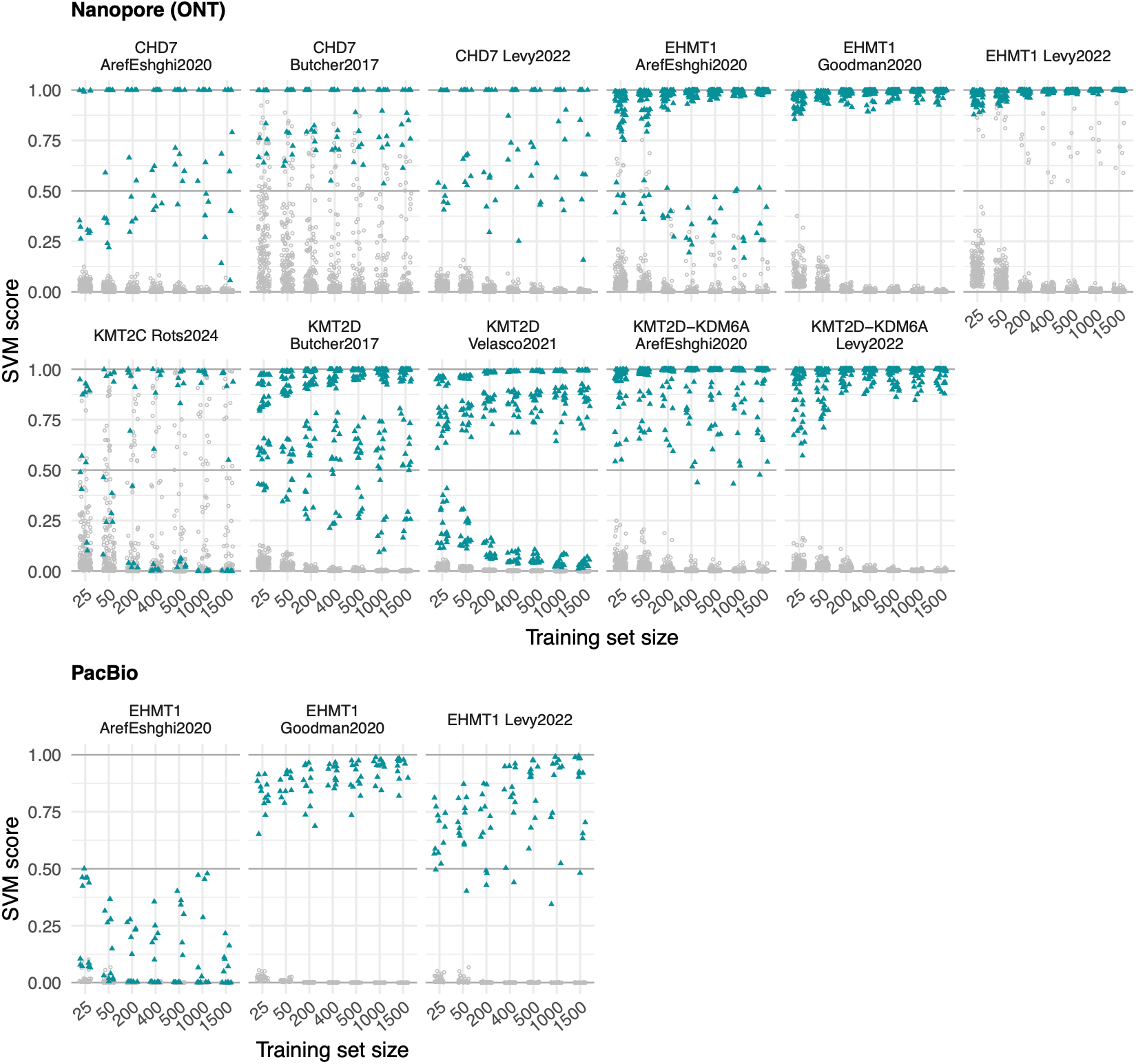
Impact of the size of the synthetic training set on the performance of array-trained DNAm classifiers applied to long-read sequencing data. Each individual is represented by six points per training set size, one for each of the six SVMs trained on different pairs of control folds. SVM score represents the probability of a sample being classified as a positive case for the specific DNAm signature. Affected individuals are shown in teal, controls in grey.

**Supplementary Figure 13.**
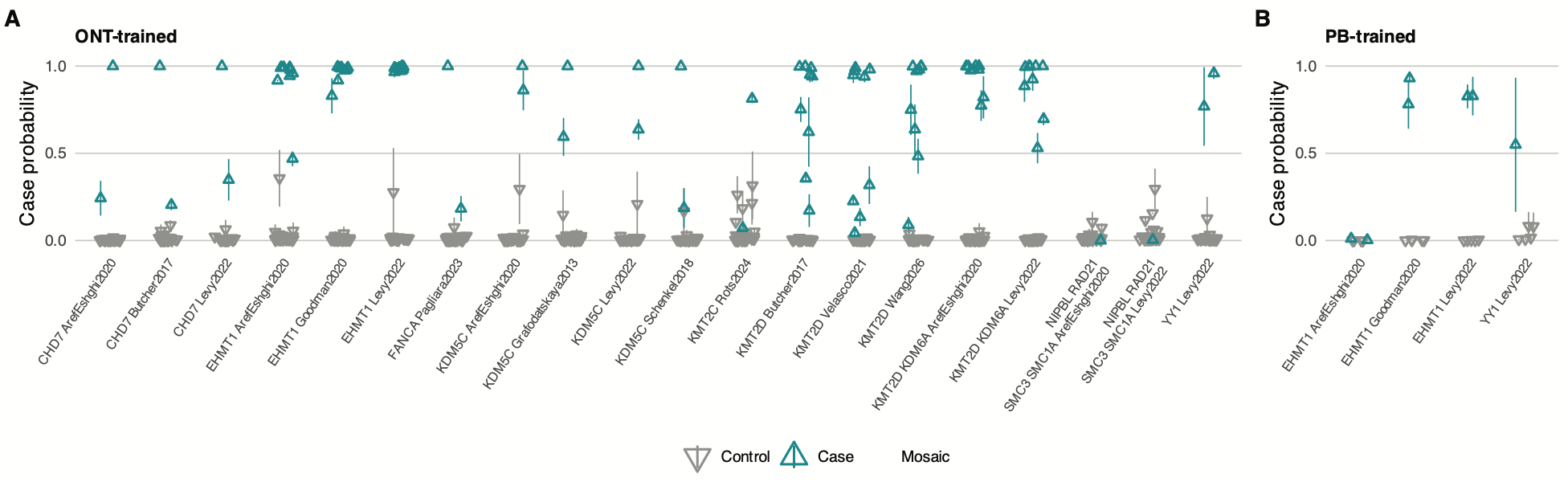
Synthetic long read epigenome-trained MethaDory classifiers identify affected individuals in matching technology A) Classification scores of ONT samples with MethaDory ONT-trained classifiers, B) Classification scores of PacBio samples with MethaDory PacBio trained classifiers.

**Supplementary Figure 14.**
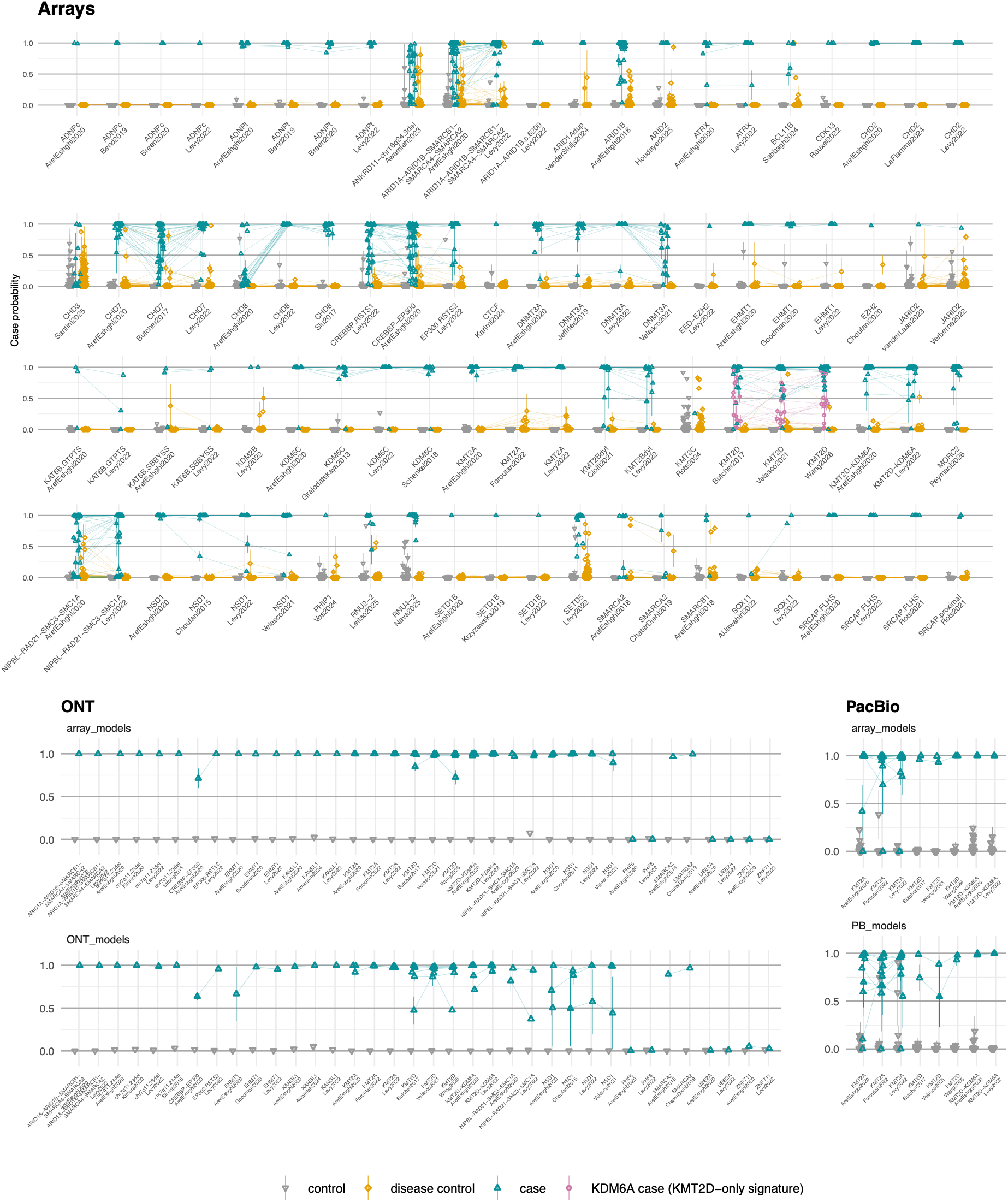
MethaDory classifiers external validation. Classification scores of samples assessed on arrays, ONT and PacBio. For ONT and PacBio both array-based MethaDory and technology-matched were run.

**Supplementary Figure 15.**
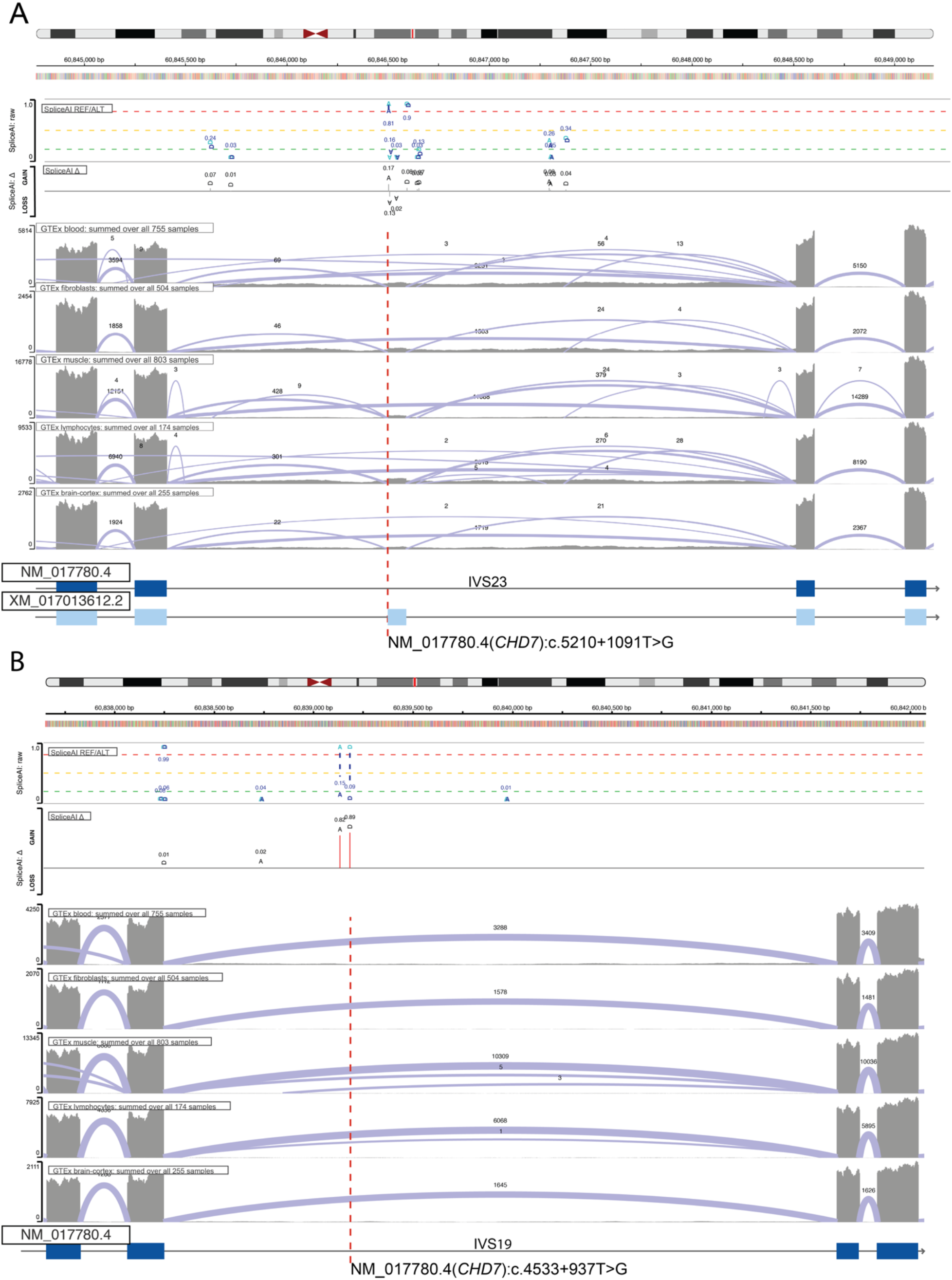
Splicing analysis of the deep intronic CHD7 variants identified by long-read genome sequencing. Predicted splicing consequences of the two deep intronic CHD7 variants identified by Oxford Nanopore long-read genome sequencing in individuals positive for the CHD7 DNAm signature by MethaDory: **A)** NM_017780.4(CHD7):c.5210+1091T>G, predicted by SpliceAI to lead to inclusion of a cryptic in-frame exon, and **B)** NM_017780.4(CHD7):c.4533+937T>G, predicted to lead to inclusion of an out-of-frame exon.

## Notes

### Competing Interest Statement

MD, DR have received travel support from Oxford Nanopore Technologies. The other authors have no conflicts of interest.

### Summary of Updates

The present update is extended with more disorders, additional classifier architecture, as well as the long-read classification, the multicentre external and prospective clinical validations, the replicability analysis, and the clinical applications of MethaDory.

